# CHARACTERIZING AND MANAGING AN EPIDEMIC: A FIRST PRINCIPLES MODEL AND A CLOSED FORM SOLUTION TO THE KERMACK AND MCKENDRICK EQUATIONS

**DOI:** 10.1101/2021.09.09.21263355

**Authors:** Ted Duclos, Tom Reichert

## Abstract

We derived a closed-form solution to the original epidemic equations formulated by Kermack and McKendrick in 1927 (1). The complete solution is validated using independently measured mobility data and accurate predictions of COVID-19 case dynamics in multiple countries. It replicates the observed phenomenology, quantitates pandemic dynamics, and provides simple analytical tools for policy makers. Of particular note, it projects that increased social containment measures shorten an epidemic and reduce the ultimate number of cases and deaths. In contrast, the widely used Susceptible–Infectious–Recovered (SIR) models, based on an approximation to Kermack and McKendrick’s original equations, project that strong containment measures *delay* the peak in daily infections, causing a longer epidemic. These projections contradict both the complete solution and the observed phenomenology in COVID-19 pandemic data. The closed-form solution elucidates that the two parameters classically used as constants in approximate SIR models cannot, in fact, be reasonably assumed to be constant in real epidemics. This prima facie failure forces the conclusion that the approximate SIR models should not be used to characterize or manage epidemics. As a replacement to the SIR models, the closed-form solution and the expressions derived from the solution form a complete set of analytical tools that can accurately diagnose the state of an epidemic and provide proper guidance for public health decision makers.

## 1. Introduction

In their seminal paper published in 1927 (1), Kermack and McKendrick developed a system of integro-differential equations for modeling epidemics. The authors made several attempts to find a closed-form solution, but ultimately were unsuccessful. At the end of their publication, in an attempt to extract a useable model from their analysis, they proposed an approximation, now known as the Susceptible–Infectious–Recovered (SIR) model (1). The equations of the SIR model were derived using the assumption that two key parameters in the original integro-differential equations could be approximated as constants.

The now widely known SIR approximation (referenced herein as the ASIR equations) used in projections of epidemic spread are said to predict that social distancing will “flatten the curve” (i.e., reduce and delay the peak of new daily cases). With cartoons of this notion highlighted in the popular media, concerns about the economic devastation associated with containment measures applied in response to COVID-19 caused many individuals and governments to advocate that social distancing measures be lifted as soon as possible (2) to shift the peak forward and truncate economic, educational, and social disruption.

As the COVID-19 pandemic swept across the world in the winter and spring of 2020, different countries applied diverse mitigation measures (3–5) in their attempts to control the spread of the virus. In so doing, these countries unintentionally conducted natural experiments on the effectiveness of differing levels of containment. Their case data provides an historically unique opportunity to compare the projections from different models using real-world data.

In this manuscript, we first derive a closed-form solution to the original equations proposed by Kermack and McKendrick (1). We verify this solution mathematically, by correlating different aspects of the model to independent data, and we accurately predict time series of case data for a sample of countries that used differing combinations and degrees of social containment measures. We call the model underlying the closed-form solution, the complete SIR, or CSIR model.

Following the derivation of the solution, we then compare epidemic dynamics predicted by the CSIR model solution and the ASIR model to the dynamics seen in countries during the COVID-19 pandemic. These comparisons show immediately that ASIR models project trends opposite to those seen in COVID-19 case data from countries, while those projected by the CSIR model solution are consonant with the real-world trends.

Inspection of the ASIR and CSIR model equations reveals that both models have two terms that represent critical elements in an epidemic and are assumed to be constant. One of these terms putatively represents the transmissibility of the epidemic entity; the other represents the behavior of the population affected. These terms differ in the two models, and the consequences of assuming one set to be constant can be determined in the other model.

Assuming the ASIR “constants” to be invariable and imposing this constraint on the CSIR model subsequently leads to implausible and unrealistic implicit assumptions regarding the dynamic behavior of the disease and the affected population. While the thus-modified CSIR model matches the ASIR projections precisely; these projections fail to match the real-world data from various countries. The contrapuntal proposition in which the CSIR model “constants” are assumed to be invariant leads to a simple variation in the ASIR model constants, which can be calculated directly from the CSIR model. The then-modified ASIR model correctly projects trends and accurately matches real-world data.

Finally, the availability of a closed-form enables the derivation of quantitative tools that can be used to manage epidemics. The details are found in the supplements to this manuscript.

## 2. Models

### 2.1 Complete susceptible infected recovered model (CSIR)

In 1927, Kermack and McKendrick (1) developed the following set of integro-differential equations for modeling epidemics (Note: We use the following equation notation, (X, SY-Z), where X is the equation number in the body, Y is the supplement (S) number and Z is the number of the equation in the supplement):

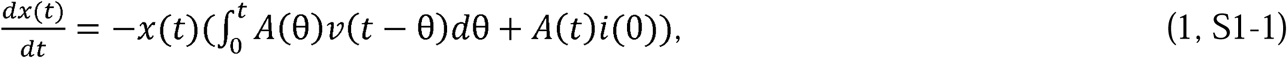

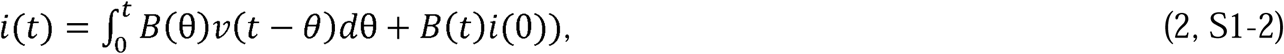

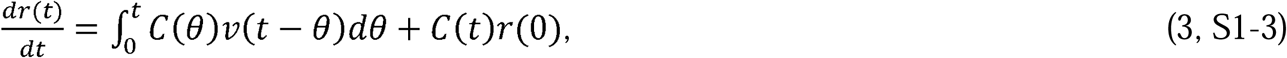

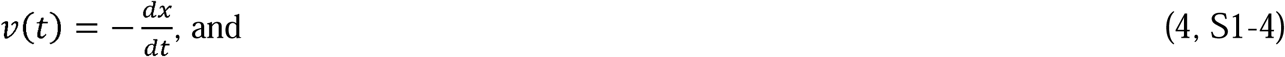

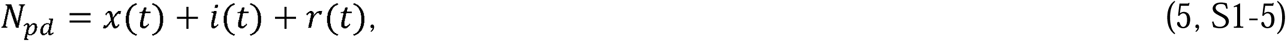

Where 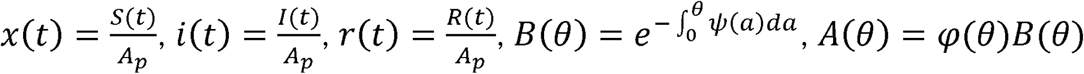, and *C(θ)* =*ψ (θ) B(θ)*. Kermack and McKendrick (1) defined *φ(θ)* as “the rate of infectivity at age *θ*” (page 703), *ψ (θ)* and as “the rate of removal” (page 703) of the infected population. We define *N*_*pd*_ as the population density; *A*_*p*_ as the area that contains the population; *S*(*t*) as the number of people susceptible to infection at time *t, I(t* as the number of people infected at time *t, R*(*t*) as the number of people recovered at time *t, N*_*p*_ as the total number of people in the population; and therefore, *N*_*pd*_*A*_*p*_ *= N*_*p*_. The variables, θ and *a*, are dummy variables for time with *θ* defined as the stage of the infection (i.e., *θ =* the time interval since the infection). We call this set of equations, developed by Kermack and McKendrick, the Complete SIR (CSIR) model.

In Supplements 1.1 and 1.2, we develop a solution to Equations 1 to 5 by first recasting them into an equivalent set of differential equations, and then solving these new differential equations starting with a first principles argument. Subsequently, in Supplement 1.3, we develop the functional forms of *φ(t), ψ(t), φ(θ)* and *ψ (θ)*. These are:

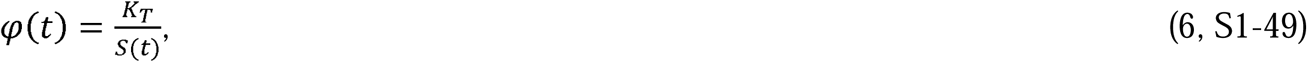

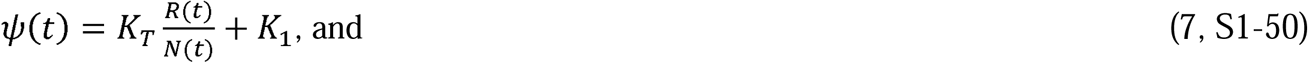

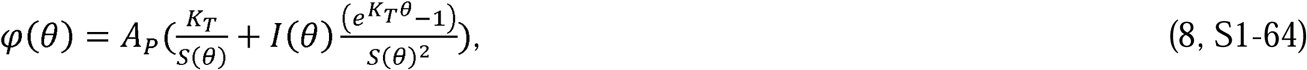

where 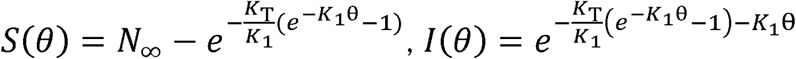, and

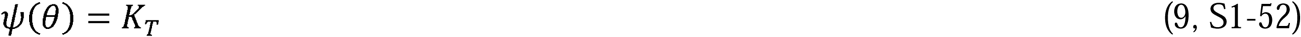

In equations 6-9, *K*_*T*_ is a constant representing the disease transmissibility with the units of people infected/(infected people x time) = 1/time, *P*_*c*_ is a constant with the units of number of infectious contacts/person representing the number of specific people an individual contacts in an infectious manner during the time from *t*_0_ to *t* (*P*_*c*_ describes the social interaction of the population), and 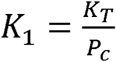 with the units of people/(infectious contacts x time) = 1/time.

Also in Supplement 1.3, using *φ(t), ψ(t), φ(θ)*, and *ψ(θ)*, we then show that the following are the functional forms of *I(t),R*(*t*), and *S*(*t*) that satisfy Equations 1 to 5:

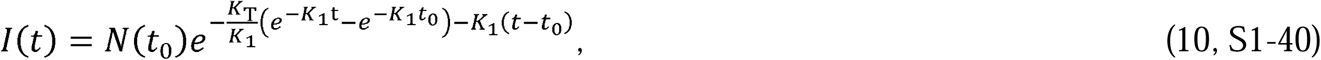

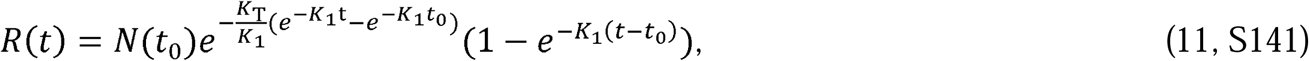

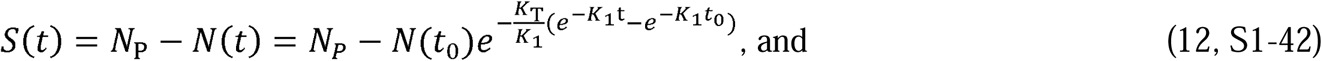

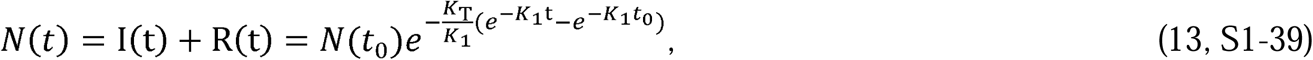

By definition, *N*(*t*) in Equation 13 is the total number of people infected since the start of the epidemic. Since 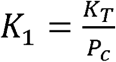, only two independent parameters, *K*_*T*_ and *P*_*c*_, which represent, respectively, the disease and the population behavior, are required to characterize the solution. Equations 10 to 13 are a closed-form solution to the CSIR model (Equations 1 to 5) proposed by Kermack and McKendrick (1); and as we show in the next section, they accurately describe the evolution of an epidemic during stages when the disease and population behavior, represented by *K*_*T*_ and *P*_*c*_, can reasonably be assumed to be constants. A more general solution to the Kermack and McKendrick CSIR equations with *K*_*T*_ and *P*_*c*_ as functions of time is derived in Supplement 3.

#### 2.1.1 Veracity of the CSIR Solution

To demonstrate the predictive capability of the CSIR solution (Equations 10 to 13), we applied it to real-world data from several countries. The first step in the process was to estimate two country-specific constants, *K*_1_ and *K*_2_. The path to these constants was found by first differentiating Equation 13 to arrive at an expression for 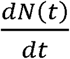, the rate at which new cases arise:

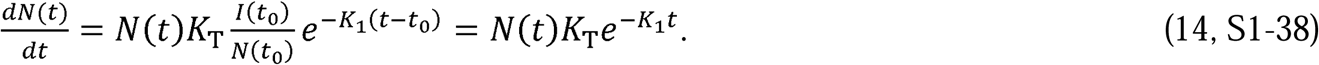

The form of Equation 14 is mathematically suggestive. A purposefully chosen combination and rearrangement of Equations 13 and 14 leads to the linear expression

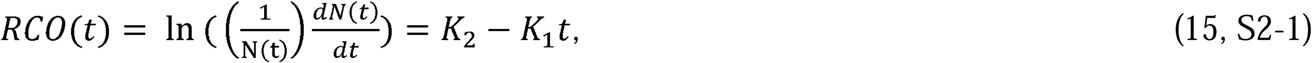

where 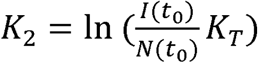 and *RCO*(*t*) means Rate of Change Operator (RCO).

We call this expression the rate of change operator because it is the rate of change of new cases scaled by the current total of cases. As we will demonstrate, Equation 15 enables the determination of the parameters *K*_1_ and *K*_2_, and consequently *K*_T_ and *P*_*c*_, for each country.

Using published data from the COVID-19 epidemic (6), we calculated and plotted the RCO time series for six different countries (Figure 1). In Figure 1 each time series has a distinctive segment where the curve appears to become a straight line. This phenomenon appears shortly after each country imposed intervention measures (indicated by arrows in the plots; the dates are listed in the caption of Figure 1). The appearance of these linear sections in every case supports the model validity and justifies the assumption that *K*_1_ and *K*_2_ may be assumed to be constant. Additionally, since 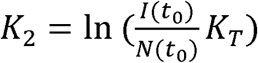 and 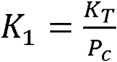, both *K*_T_ and *P*_c_ can also be modelled as constants for the periods of time during which the RCO series remains linear.

**Figure 1.**
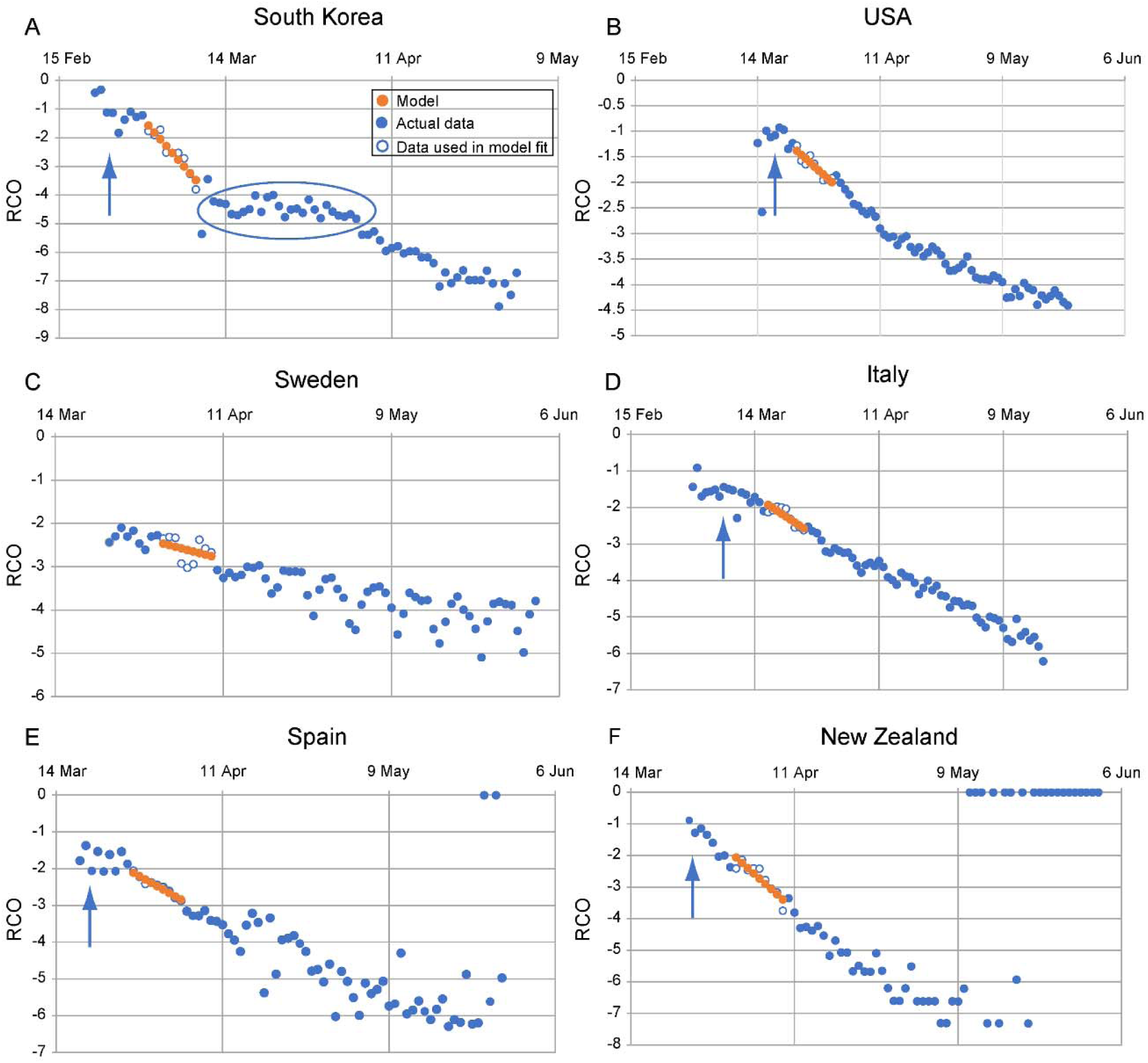
Rate of change operator (RCO) curves for COVID-19 cases in various countries. An epidemic can be described by a piecewise linear model using the RCO (Equation 15). A short segment of orange dots in each graph is a linear fit to the corresponding points (blue/white circles) in the observed data. The slopes and initial points of these dotted-line segments are the values of *K*_1_ and *K*_2_ respectively which are tabulated in Table1. In some countries, RCO curves changed markedly soon after the date containment measures were implemented (arrows): **A)** South Korea, February 21; (the oval highlights a departure of the observed data from the RCO slope, indicating failures in, or relaxations of, social distancing); **B)** USA, March 16; **C)** Sweden did not implement any specific containment measures, so the model calibration was begun on April 1, the date when the slope of the RCO curve first became steady. **D)** Italy, March 8; **E)** Spain, March 14; **F)** New Zealand, March 25. All dates are in 2020.

Using Equation 15, the parameters *K*_1_ and *K*_2_ for each country were determined by finding the slopes and intercepts of lines fitted to short, early portions of the straight segments of the RCO time series. These early portions comprised nine data points each (see Table 1 for their date ranges); and Table 1 displays the *K*_1_ and *K*_2_ values derived for each country.

**Table 1.**
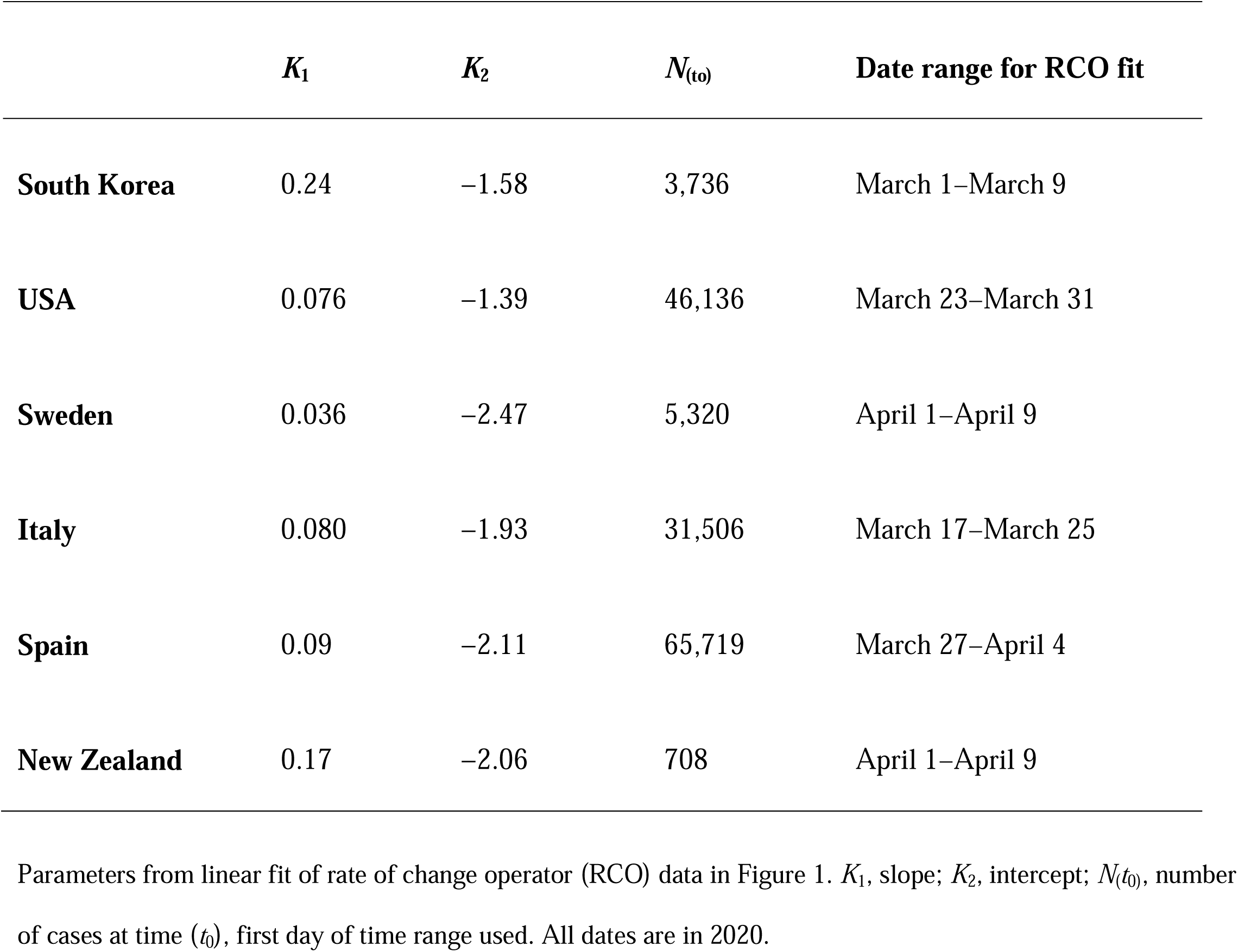
Social containment parameters used to model total cases and new daily cases of infection for different countries (6).

| | $K_1$ | $K_2$ | $N_{(t_0)}$ | Date range for RCO fit |
| --- | --- | --- | --- | --- |
| **South Korea** | 0.24 | -1.58 | 3,736 | March 1–March 9 |
| **USA** | 0.076 | -1.39 | 46,136 | March 23–March 31 |
| **Sweden** | 0.036 | -2.47 | 5,320 | April 1–April 9 |
| **Italy** | 0.080 | -1.93 | 31,506 | March 17–March 25 |
| **Spain** | 0.09 | -2.11 | 65,719 | March 27–April 4 |
| **New Zealand** | 0.17 | -2.06 | 708 | April 1–April 9 |

Using the values of *K*_1_ and *K*_2_ in Table 1 in Equation 13 we then predicted the course of daily total cases (Figure 2) for the six countries. These predictions matched the actual time series of the daily total cases with an *R*^*2*^ > 0.97 in each of the six countries for the 45 days following the date containment measures were introduced. The CSIR solution also predicted daily new cases using Equation 14 (Figure 3) for the six countries for the same 45 days with an *R*^*2*^ range of 0.29 to 0.90. As seen in the figure, the predicted peak of new cases was close to the observed peak for all countries.

**Figure 2.**
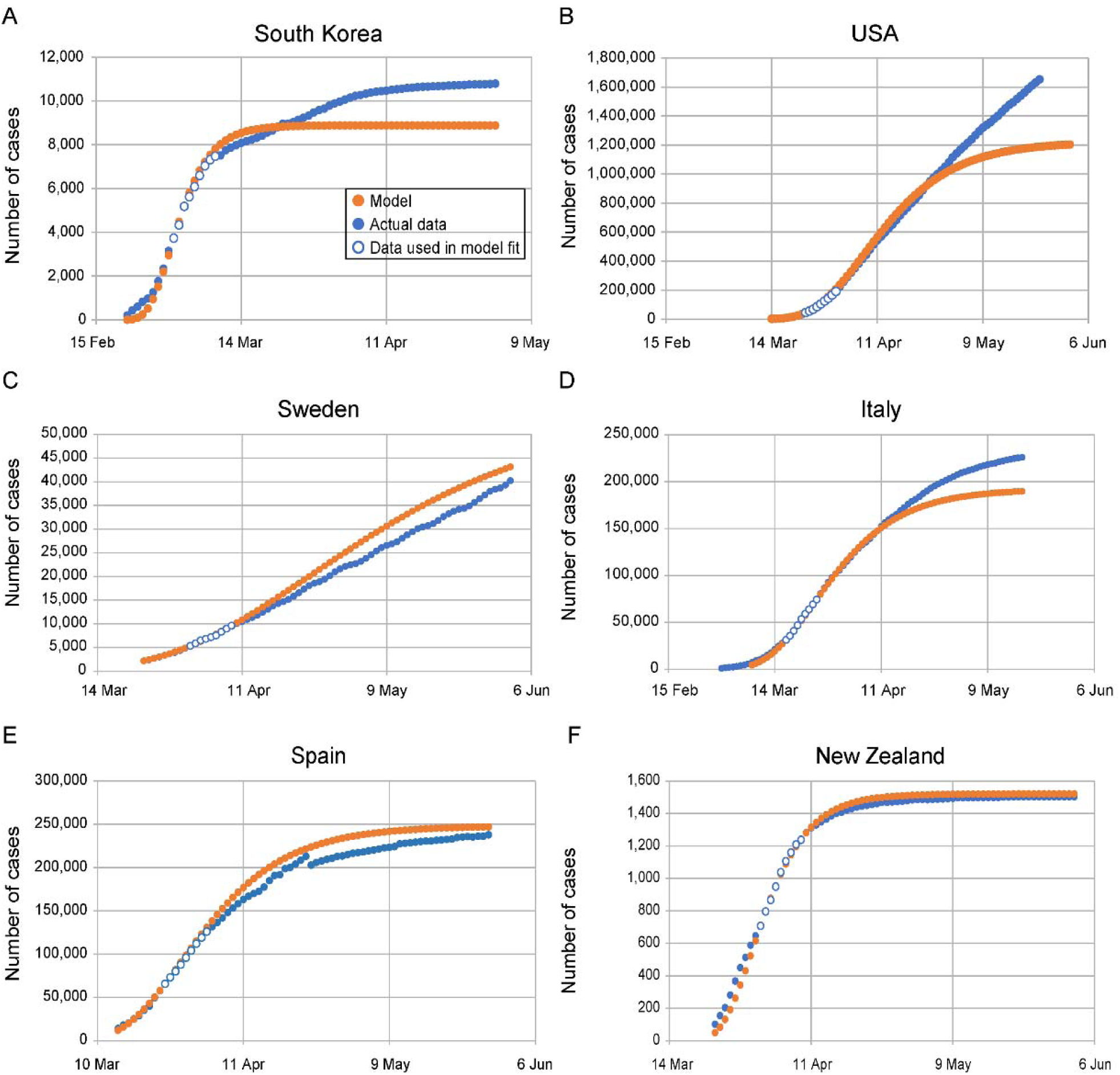
Complete SIR (CSIR) model predictions for daily total case counts. **A)** South Korea; **B)** USA; **C)** Sweden; **D)** Italy; **E)** Spain; and **F)** New Zealand. Dots are daily data points observed from (white-center and all blue) or calculated (orange) for each country. The CSIR model was calibrated using data from the date ranges listed in Table 1 (white-center blue dots). *R*^2^ > 0.97 for the model fit for all countries for the 45 days after the containment measures were implemented: South Korea, February 21-April 4; USA, March 16-April 30; Italy, March 8–April 22; Spain, March 14-April 28; New Zealand, March 25-May 9. Sweden did not implement any specific containment measures, so the dates used were March 23-May7. The deviation of the model from the data in the USA, panel (**B**), after April is elucidated in Supplement 5. All dates are in 2020.

**Figure 3.**
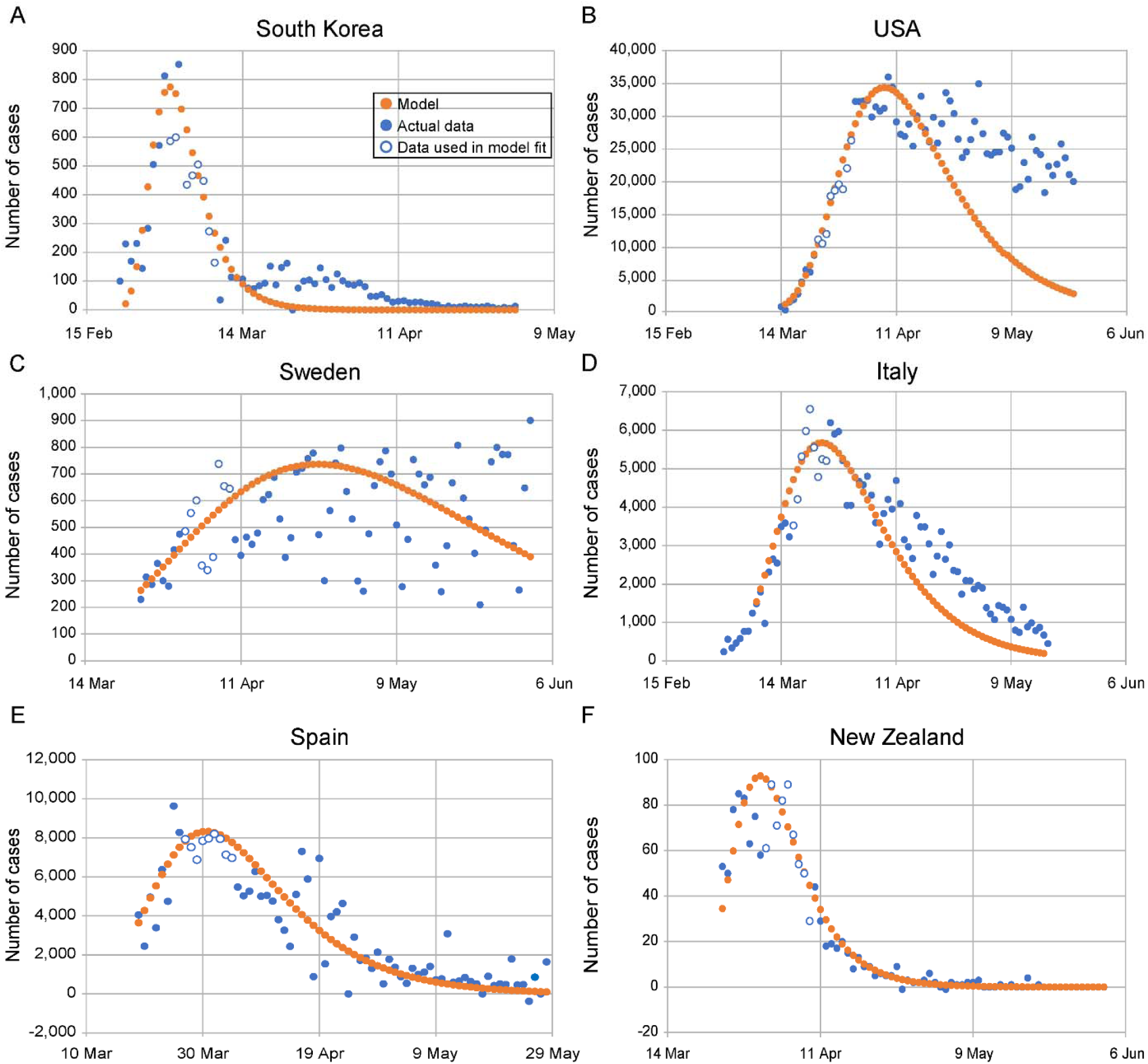
Complete SIR (CSIR) model predictions for number of new daily cases. **A)** South Korea, *R*^2^ = 0.86; **B)** USA, *R*^2^ = 0.83; **C)** Sweden, *R*^2^ = 0.29; **D)** Italy, *R*^2^ = 0.69; **E)** Spain, *R*^2^ = 0.65; and **F)** New Zealand, *R*^2^ = 0.90. The orange dotted line is the model in all panels. The all-blue and white-center blue dots are data points, daily observations from each country. The white-center blue points are used to determine model parameters. *R*^2^ values are between the model and the data, across countries for the 45 days after containment measures were imposed. All dates are in 2020.

It is important to emphasize that the CSIR predictions in Figures 2 and 3 are *not* fits to the full-length of the data shown. Rather, the two constants, *K*_1_ and *K*_2_ were estimated using only a short, linear, nine-point portion of the epidemic data starting between 7 to 14 days after the imposition of containment measures. These constants were then used to project the data following these nine points.

In an additional demonstration of the veracity of the CSIR solution, we tested the assumption that *K*_T_ is a property of the disease, and therefore, should be the same for each country. Equation S2-5 shows that the model parameters, expressed in a purposefully constructed function, *F(N(t))*, should be linearly proportional to time with a constant of proportionality or slope equal to −*K*_*T*_. As illustrated in Figure 4 and explained in more detail in Supplement 2, the fit of Equation S2-5 using the population density data from Table 2 has an *R*^*2*^ = 0.956 and a slope of −0.26 (the slope is equal to −*K*_*T*_). This excellent correlation confirms that *K*_*T*_ can confidently be assumed to be the same for all countries (=0.26), a constant, and plausibly, a property of the disease.

**Table 2.**
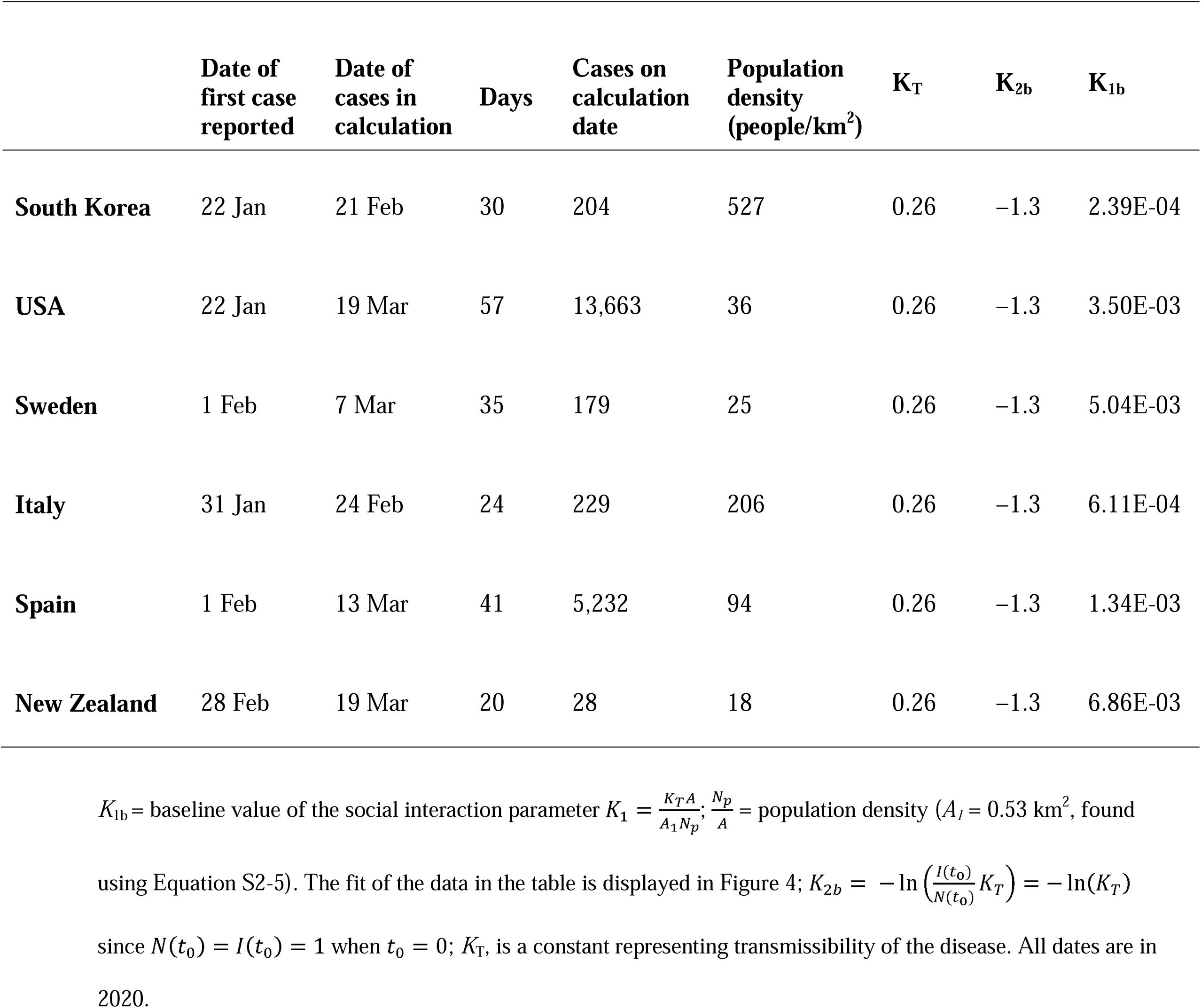
Initial COVID-19 pandemic data and social interaction parameters for various countries ((6), case and date data; (7), population density data)

| | Date of first case reported | Date of cases in calculation | Days | Cases on calculation date | Population density (people/km<sup>2</sup>) | $K_T$ | $K_{2b}$ | $K_{1b}$ |
| --- | --- | --- | --- | --- | --- | --- | --- | --- |
| **South Korea** | 22 Jan | 21 Feb | 30 | 204 | 527 | 0.26 | -1.3 | 2.39E-04 |
| **USA** | 22 Jan | 19 Mar | 57 | 13,663 | 36 | 0.26 | -1.3 | 3.50E-03 |
| **Sweden** | 1 Feb | 7 Mar | 35 | 179 | 25 | 0.26 | -1.3 | 5.04E-03 |
| **Italy** | 31 Jan | 24 Feb | 24 | 229 | 206 | 0.26 | -1.3 | 6.11E-04 |
| **Spain** | 1 Feb | 13 Mar | 41 | 5,232 | 94 | 0.26 | -1.3 | 1.34E-03 |
| **New Zealand** | 28 Feb | 19 Mar | 20 | 28 | 18 | 0.26 | -1.3 | 6.86E-03 |

$K_{1b}$ = baseline value of the social interaction parameter $K_1 = \frac{K_T A}{A_1 N_p}$ ; $\frac{N_p}{A}$ = population density ( $A_l = 0.53 \text{ km}^2$ , found
using Equation S2-5). The fit of the data in the table is displayed in Figure 4; $K_{2b} = -\ln\left(\frac{I(t_0)}{N(t_0)} K_T\right) = -\ln(K_T)$
since $N(t_0) = I(t_0) = 1$ when $t_0 = 0$ ; $K_T$ , is a constant representing transmissibility of the disease. All dates are in 2020.

**Figure 4.**
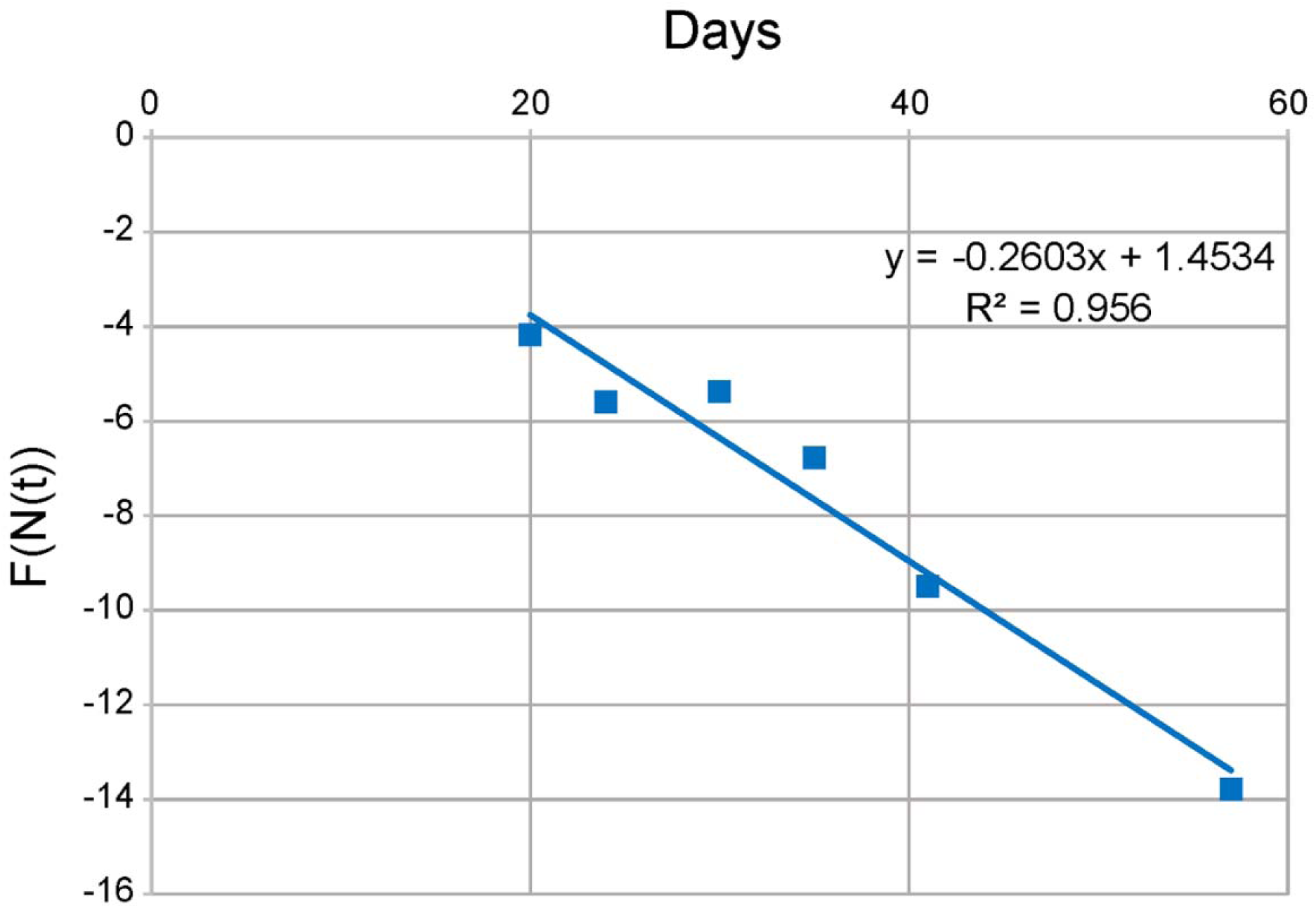
Verification that *K*_*T*_ is the same for all countries. The data from Table 2 is plotted using Equation S2-5 and *A*_*1*_ = 0.48 *km2*. Each data point corresponds to a different country. The value of *K*_*T*_ is the negative of the slope of the line, and *K*_*T*_ is closely approximated everywhere by *K*_*T*_ ≈ 0.256.

A third illustration of veracity arises from the ability to correlate independently sourced mobility data to the RCO from the CSIR model. Based on Equations S2-1 and S3-8 derived in Supplements 2 and 3, if the CSIR model solution is correct, then the integral of this mobility data should correlate linearly with the measured RCO. Mobility data, available from Google (8), are a measure of the difference between the amount of time people stayed at home (the Residential Mobility Measure or RMM) during the period modelled and a baseline measured for 5 weeks starting January 3, 2020. Figure 5 shows that, as the CSIR model solution predicts, for each country considered, the integral of the RMM and the RCO are linearly correlated to a high degree.

**Figure 5.**
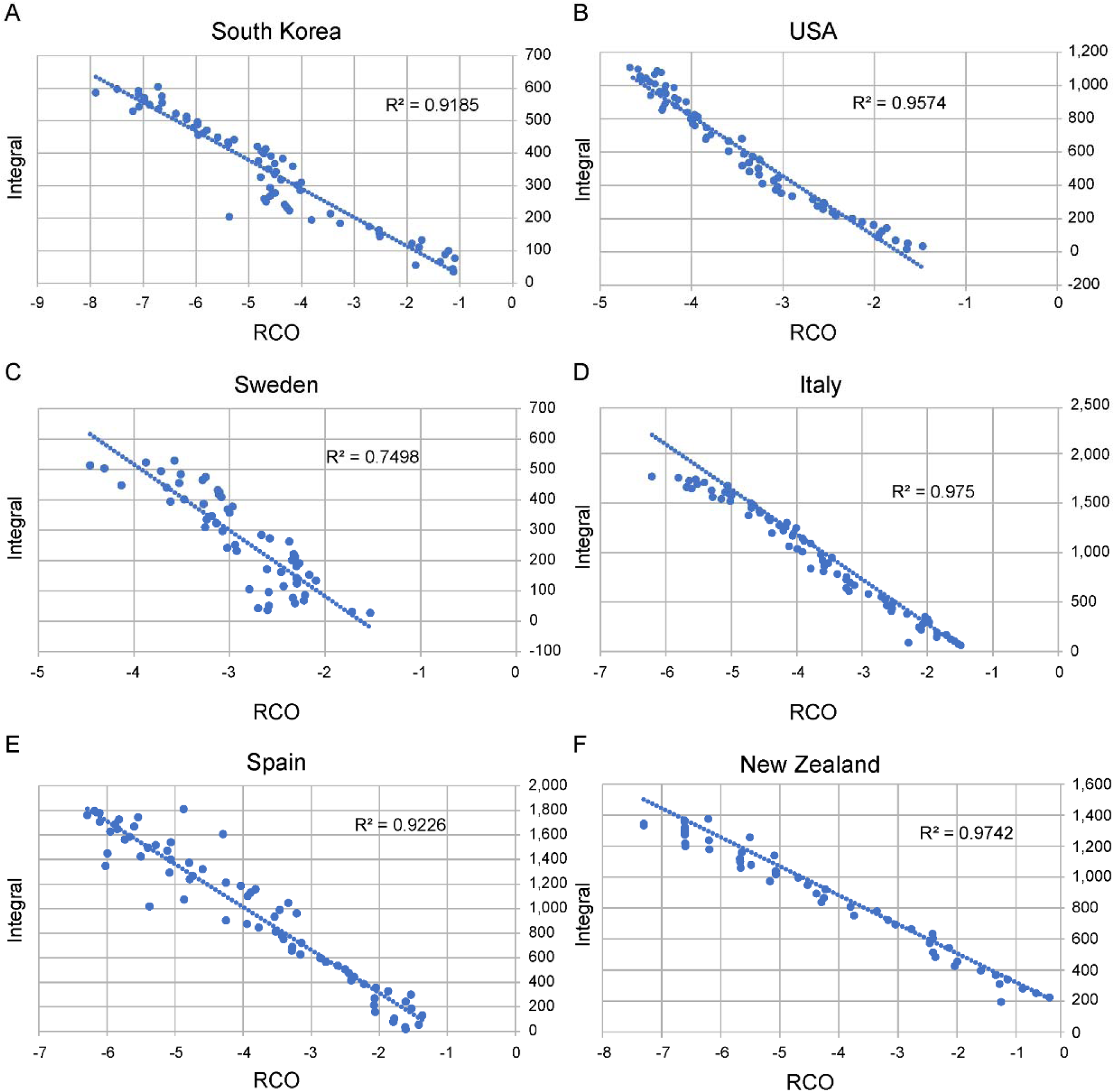
Correlations between the daily value of the rate of change operator (RCO) and the integral of the Google Residential Mobility Measure (RMM) (8). **A)** South Korea (date range, February 23 to April 23); **B)** USA (March 25 to May 31); **C)** Sweden (March 5 to May 5); **D)** Italy (March 25 to May 31); **E)** Spain (March 25 to May 31); and **F)** New Zealand (March 21 to April 22). All dates are in 2020.

As orthogonal correlative support for the veracity of the CSIR solution, we note that other authors (9) have observed that the daily COVID-19 case data from many countries can be fit with a Gompertz model. Since Equation 13 is also a Gompertz equation, those observations support the assertion that Equations 10 to 13 properly model epidemics.

Taken together, these results demonstrate that the solution to the CSIR model correctly characterizes epidemic dynamics from multiple countries in a unified way. They also demonstrate that the CSIR model developed by Kermack and McKendrick (1) nearly 100 years ago, described by Equations 1 to 5, was correct and accurately captures the dynamics of epidemics.

### 2.2 The Approximate SIR (ASIR) model

Kermack and McKendrick attempted to find an analytical solution for the CSIR equations, but they were not successful. As an alternative, they proposed the following approximation to the CSIR equations:

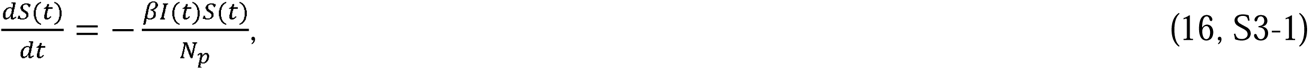

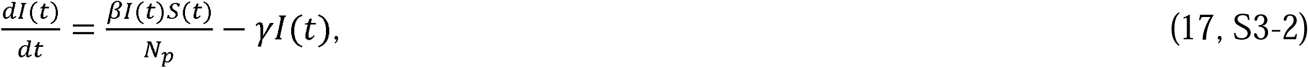

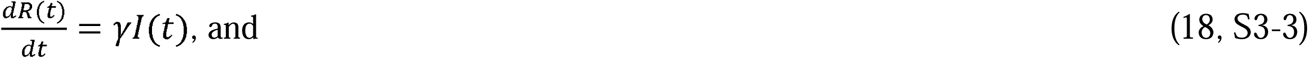

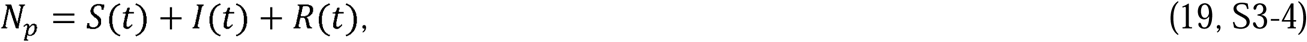

where *β* = rate of contact and transmission, *γ* = rate of recoveries = 1/*t*_*r*_, and *t*_*r*_ = time of infectiousness.

These equations are the well-known “SIR” model equations, which we have previously designated as the approximate SIR (ASIR) model equations. They can be derived from Equations 1 to 5 by assuming that the parameters 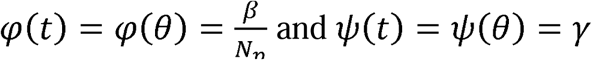 are constants. The equations of the ASIR model and variants (SEIR, MSEIR, etc.) have been used for decades in attempts to quantitatively and qualitatively model epidemics.

Since *β* =*φN*_*p*_ and *φ* was defined by Kermack and McKendrick (1) as “the rate of infectivity at ageθ” (page 703), *β* has generally been interpreted as a measure of social containment in the at-risk population. Modelers have assumed that a lower *β* indicates higher social containment. Likewise, since *γ* = *ψ* and Kermack and McKendrick defined *ψ* as “the rate of removal” (page 703) of infected persons to a recovered state or death, *γ* is generally interpreted as a measure of persistence of infectiousness, a parameter associated with the agent of the disease; a lower *γ* has been assumed to represent longer lasting disease.

A simulation of the ASIR model, depicted in Figures 6A and B, shows that the model projects that an increase in social containment (decreasing *β*) causes a later end to the epidemic and a lower and progressively later peak in cases per day. In contrast, a plot of the CSIR model solution in Figures 6C and D exhibits the opposite phenomenology: an increase in social containment (higher *K*_1_) causes an *earlier* end to the epidemic and a lower and progressively *earlier* peak in cases per day. As social containment measures increase, the positions of the peak in new cases per day move in *opposite* directions for the two models.

**Figure 6.**
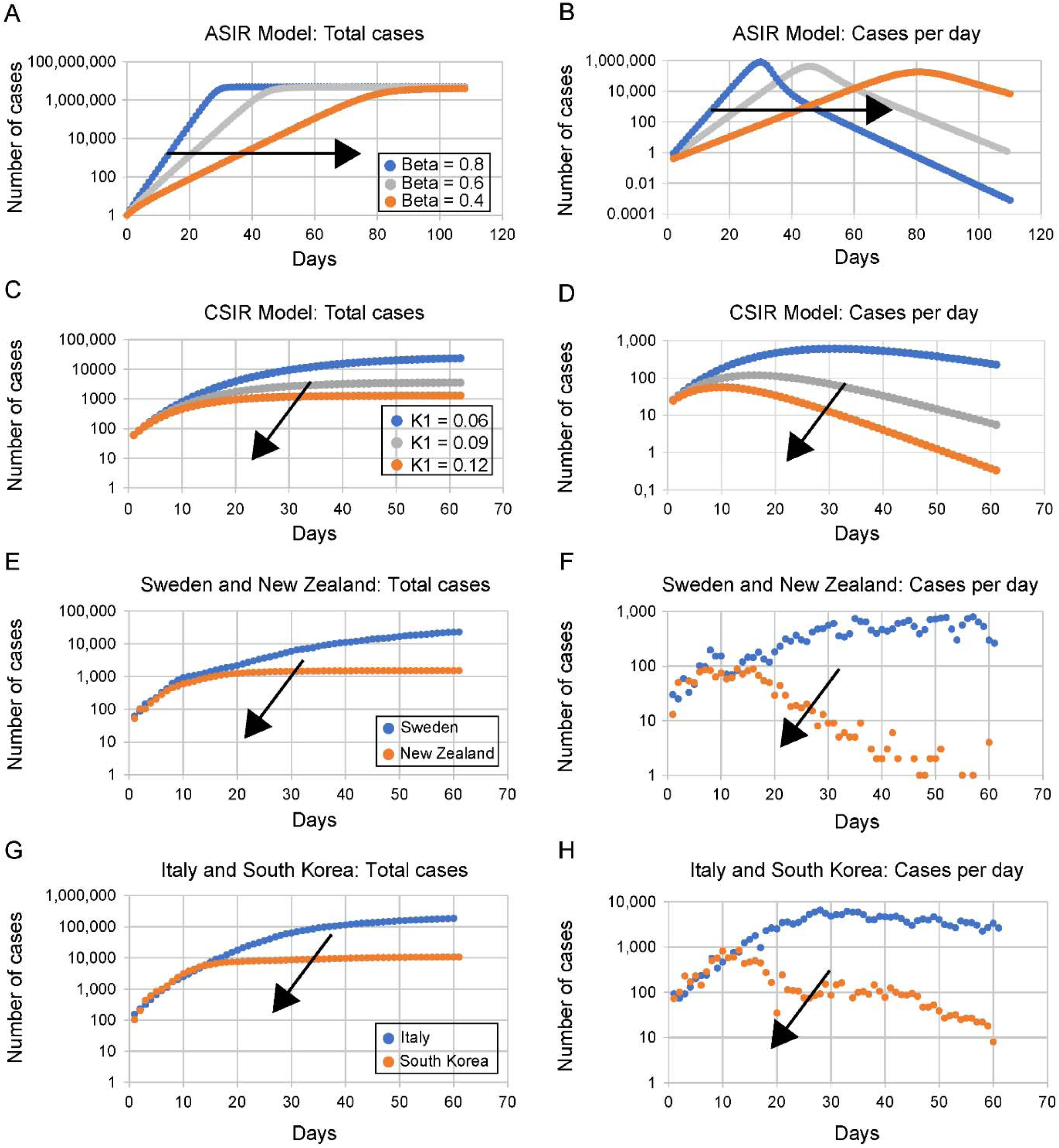
Comparisons of predictions of the approximate SIR (ASIR) and complete SIR (CSIR) models with observed data from four countries. ***Note: Containment measures increase in all panels from blue to grey to orange dot curves. The arrow on each graph indicates the direction of more social distancing***. **ASIR model trend predictions: (A)** Total cases; **(B)** Daily new cases. Rate of contact and transmission (β) decreases with increasing social distancing (from blue to grey to orange curves). Rate of recoveries (γ) = 0.2 for both sets of plots. As β decreases, the daily total of cases increases more slowly and plateaus later (**A**). Daily new infections project to later, but only slightly lower peaks (**B**). **CSIR model trend projections: (C)** Total cases; **(D)** Daily new cases. As containment measures increase (higher *K*_1_, blue to grey to orange curves), Equation 13 projects that total cases will rise to lower levels; and reach these levels earlier **(C)**. Similarly, Equation 14 projects that new daily cases will peak earlier to lower values with increasing containment **(D)**. *K*_*T*_ = 0.2 for plots **(C)** and **(D)**. **Data reported from different countries during the COVID-19 pandemic**. The remaining graphs contain data from pairs of countries with differing containment measures referenced to a day when each member of the pair had nearly equal numbers of new cases. **(E)** Total cases in Sweden (no containment measures, blue) and New Zealand (strict containment, orange).**(F)** Daily new cases in Sweden (blue) and New Zealand (orange).**(G)** Total cases in Italy (loose containment measures, blue) and South Korea (strict containment, orange). **(H)** Daily new cases in Italy (blue) and South Korea (orange). The trends in the observed data, panels (**E – H)**, are the opposite of those exhibited by the ASIR model for increasing containment (decreasing β) in panels (**A, B**). The ASIR model trends in (**A**) and (**B**) have completely different shapes; and vary with increasing containment in an opposite sense to those in the country data. The CSIR model trends in **(C)** and **(D)** are highly similar to those in the country data **(E – H)**.

We can also mathematically compare the trends projected by the ASIR model with trends predicted by the CSIR model using the following expression derived from the solution to the CSIR model (see Supplement 4 for detail):

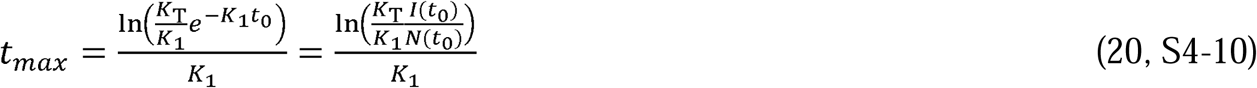

As can be deduced from Equation 20, in contrast to the ASIR solution plot in Figure 6B, the CSIR model solution mathematically predicts that the time of the peak in daily cases will occur earlier with increased social containment (i.e., higher *K*_1_).

#### 2.2.1 Testing the ASIR Model

Although we have already shown graphically and mathematically that the ASIR projections contradict the CSIR solution, the true test of a model is whether it predicts the trends present in actual data. It is fortuitous, then, that the progression of the COVID-19 pandemic has been well documented in multiple countries which took different paths while attempting to contain the spread of the virus. This dataset affords the opportunity to test the veracity of the trends predicted by both models against actual data.

In plots E to H in Figure 6, we compare the ASIR and CSIR projected trends to COVID-19 pandemic case data (6) in Sweden and New Zealand for total cases and for daily new cases (Figure 6E and F), and in South Korea and Italy (Figure 6G and H). These pairs of countries have comparable population densities but implemented mitigation measures with different intensities (3–5, 9). New Zealand and South Korea introduced stronger containment measures much earlier than Italy and Sweden.

In support of the CSIR solution and in contradistinction to the ASIR model, the country data in Figures 6E–H show that stronger containment measures are associated with an *earlier* levelling off at a *lower* total number of cases and an *earlier* and *lower* peak in new infections. Other authors (11), too, have noted that the peak of cases in countries with stronger containment measures occurred earlier than in countries with weaker measures.

Trends in both peak position and height demonstrate that the ASIR model is not merely inaccurate, a tolerable trait in an approximation, but projects epidemic data to trend in the *opposite* direction to the reported data. The ASIR model both contradicts the solution to the Kermack and McKendrick (1) original equations (ie. the CSIR model solution) and fails the simplest test of model veracity: the projection of qualitative trends.

A related observation is that the projected reduction in peak height of daily cases is greater in the CSIR model than in the ASIR model. This implies that the true impact of increased containment measures, and the concomitant decreased social interaction, is understated by ASIR models.

### 2.3 Reinterpreting the ASIR Approximation

As seen in the preceding section, though both *β* (in the ASIR model) and *P*_*c*_ (in the CSIR model) are posited to represent social interaction, the trend in the movement of the daily cases peak with decreasing *β* (less social interaction) in the ASIR model is opposite to that with decreasing *P*_*c*_ (also less social interaction) in the CSIR solution (As a reminder: an increase in *K*_1_ is the same as a decrease in *P*_*c*_). Since only the CSIR solution reproduces the trends and the values of reported epidemic data, it seems likely that the nature and implications of the ASIR assumptions may not be sufficiently understood.

Using several derived relationships from and simulations of both the ASIR model and the CSIR solution, we will now illustrate that, indeed, previously unknown implausible implications arise from the assumption that *φ*(*t*) and *ψ*(*t*) are constants in the ASIR derivation. To begin, we note that when *φ*(*t*) and *ψ*(*t*) are assumed to be the constants *φ* and *ψ*, then, by their definitions, *β* and *γ* are also constants.

As explained in Supplement 3, when *φ*(*t*) and *ψ*(*t*) are assumed to be the constants *φ* and *ψ, K*_*T*_(*t*) and *P*_*c*_(*t*) are forced to vary in the following manner:

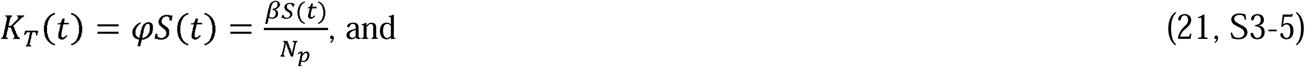

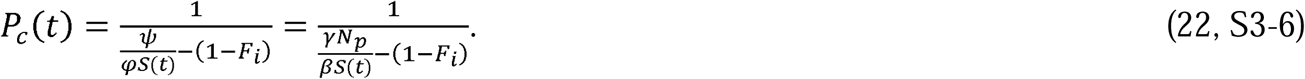

From Equation 21 we see that *φ*(*t*) can only remain constant if *K*_*T*_(*t*) decreases in direct proportion to the decreasing size of the susceptible population, *S*(*t*). This is implausible on its face because, as discussed in the derivation of the CSIR solution, *K*_*T*_(*t*) is solely a function of the disease agent and thus, is likely a constant for a substantial time at the beginning of the epidemic; at least until the disease agent itself is modified by mutation or selection.

Kermack and McKendrick themselves, in their introduction, ((1), pp. 702), also note that it is implausible to assume that disease transmissibility will *decrease* as the disease spreads.Furthermore, even if transmission were to decrease over time within the infected population, it is improbable that this decrease will occur in a fixed linear proportion to the remaining number of susceptible people as required by Equation 21. Thus, the assumption within the ASIR model that *φ* can be modelled as a constant requires an implausible additional assumption.

It is not possible to state how *P*_*c*_(*t*) must vary to maintain *ψ* as a constant by merely inspecting Equation 22. We can, however, elucidate the behavior of *P*_*c*_(*t*) required by Equation 22 by plotting the time series of Equation 22 and then deriving our conclusions from inspection of this plot. This requires a CSIR model solution with time varying *P*_*c*_, therefore, we rederived the CSIR solution (see Supplement 3) with time varying *K*_*T*_(*t*) and *P*_*c*_(*t*) to find relationships for *N*(*t*) and 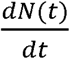 as functions of *K*_*T*_(*t*) and *P*_*c*_(*t*) (Equations S3-7 and S3-8). The time series of both *N*(*t*) and 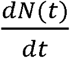 were then simulated using an Euler approximation of Equations S3-7 and S3-8 with a time step of 0.1 day. Using the same values of *β* and *γ* used in Figure 6A and B, Equations 21 and 22 were then used to determine the values of *K*_*T*_(*t*) and *P*_*c*_(*t*) employed in the simulation.

The purpose of this simulation was first to demonstrate that imposing the conditions of Equations 21 and 22 on *K*_*T*_(*t*) and *P*_*c*_(*t*) in the CSIR solution causes the CSIR model to produce the same results as the ASIR approximation. The second purpose was to determine and demonstrate the actual temporal behavior the ASIR approximation imposes on both *K*_*T*_(*t*) and *P*_*c*_(*t*).

The time series plots of the simulation of both *N*(*t*) (cases) and 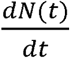 (cases per day) appear in Figure 7. The close approximation of the CSIR and the ASIR curves in Figure 7 demonstrates that the ASIR model is, indeed, a subset of the CSIR solution when the constraints of Equations 21 and 22 are applied to *K*_*T*_(*t*) and *P*_*c*_(*t*).

**Figure 7.**
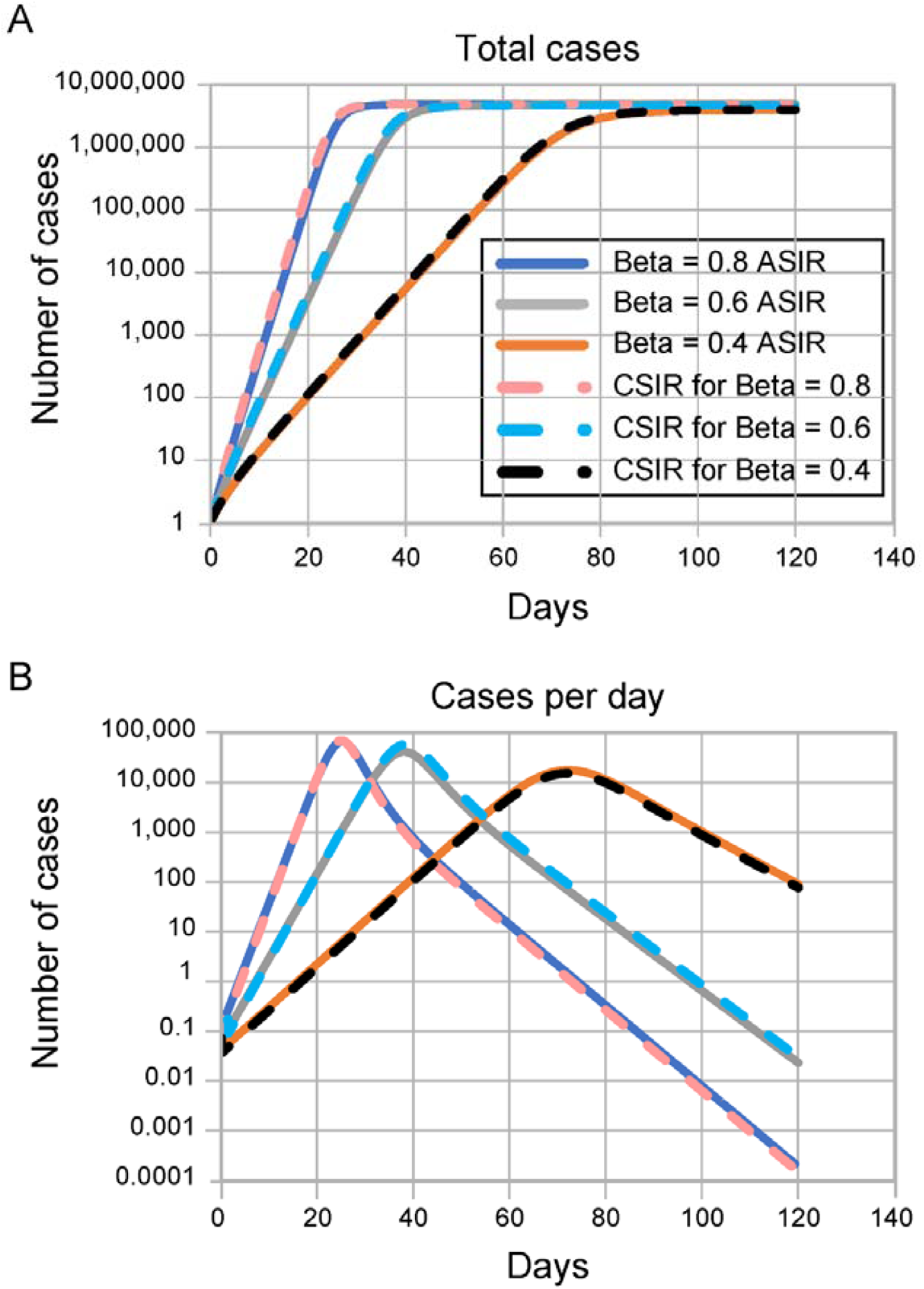
Demonstration that the CSIR model can be modified to produce the ASIR model results. Both plots contain 6 lines. For the same β and γ, both the ASIR and the CSIR simulations overlay each other. The CSIR simulations were produced by imposing the criteria in Equations S3-5 and S3-6, connecting the ASIR and CSIR constants. This plot demonstrates that the ASIR approximation provides the same result as the CSIR solution provided the constraints of these equations are imposed. γ is 0.2 for all plots. These plots are the same as the ASIR plots in figure 6A and 6B on a log scale. X axis is days.

In Figure 8 we plotted the time-associated variation of *K*_*T*_(*t*) and *P*_*c*_(*t*) that are necessary to create the CSIR curve in Figure 7. The figure shows that the constraints imposed by the ASIR model compel the acceptance of unlikely consequences; namely, *K*_*T*_(*t*) decreases with time and *P*_*c*_(*t*) must behave in an unrealistic manner.

**Figure 8.**
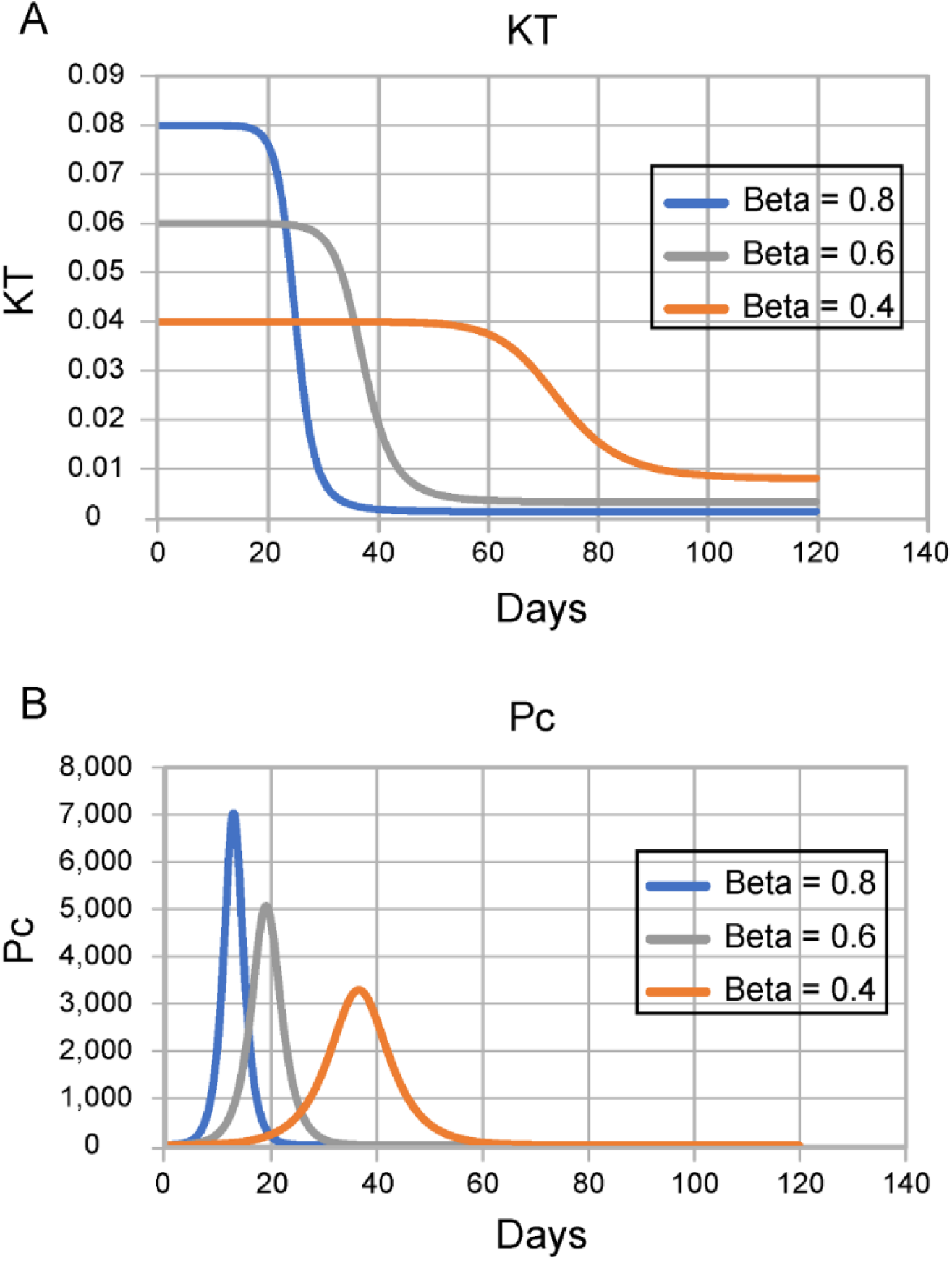
Time series for creating CSIR curves. These graphs show how Pc and KT are forced to vary within the CSIR simulation shown in Figure 7 when the constraints of Equations S3-5 and S3-6 (constant ϕ and ψ) are imposed. γ is 0.2 for all plots. X-axis is days.

While the unlikely behavior of *K*_*T*_(*t*) required by the assumptions in the ASIR model is easily deduced from Equation 21, the plot in Figure 8B explicates the complex behavior of *P*_*c*_(*t*) required by these same assumptions. When the ASIR approximation is applied, the consequent, implicit assumption is that, early in the epidemic, the population increases its contacts (rising *P*_*c*_(*t*)), and then suddenly and symmetrically (in time), reverses course and reduces the number of contacts. At each value of *β*, this up and down spike in contacts (Figure 8B) precedes a plunge in the value of *K*_*T*_(*t*) (Figure 8A), and the steep decline is immediately followed by the peak in daily cases (Figure 7B), tailing to the eventual end of the epidemic.

These implied consequences of a constant *β* in the ASIR model make the clear points that a constant *β* does *not* represent constant social interaction; and a higher *β* does *not* represent a consistently higher level of social interaction. Also, a symmetric spike in social interaction (*P*_*c*_(*t*)), higher and earlier, proportional to the value of *β*, followed by an immediate collapse in transmissibility (*K*_*T*_(*t*)), is simply unfathomable in reality.

Of note in Figure 8 is that when *β* is higher, *P*_*c*_(*t*) becomes lower earlier in the simulation and therefore the peak in Figure 7B occurs earlier. The interpretation of this observation is critical to understanding the ASIR model: Higher values of *β* in the ASIR model do represent a larger *initial* increase in social interaction. However, they also represent a subsequently earlier imposition of high containment measures because the spike in *P*_*c*_(*t*) ends earlier when *β* is higher. This earlier containment, not the initial increase in social interaction, induces the early peak in new daily cases in the ASIR model and ends the simulated epidemic earlier. In contrast to the conventional interpretation, the ASIR model does not project that higher social interaction will end the epidemic sooner.

The consequences of the approximations in the ASIR model become even more clear when we make manifest the time varying nature of *φ*(*t*) and *ψ*(*t*) (and therefore of *β* and *γ*) required when the quantities *K*_*T*_ and *P*_*c*_ are held constant. Similar to the preceding analysis, we explored these consequences using simulations of the CSIR solution and the ASIR model. Since the CSIR solution predicts the country data well, we also compared the simulation results to two of the country results (Italy and New Zealand).

As a first step, we used the values of *K*_1_ and *K*_2_ in Table 1 to derive values of *K*_1_ and *K*_*T*_ for Italy and New Zealand and used the CSIR solution to project the results. We then simulated the ASIR model with the assumption that the values of *φ* and *ψ* (and therefore of *β* and *γ*) were constant and equal to the values of *K*_1_ and *K*_*T*_ used in the CSIR solution. The results of both the CSIR and ASIR simulations are plotted in Figure 9, along with the country data. As can be easily seen, the CSIR model accurately models the country data and the ASIR model does not.

**Figure 9.**
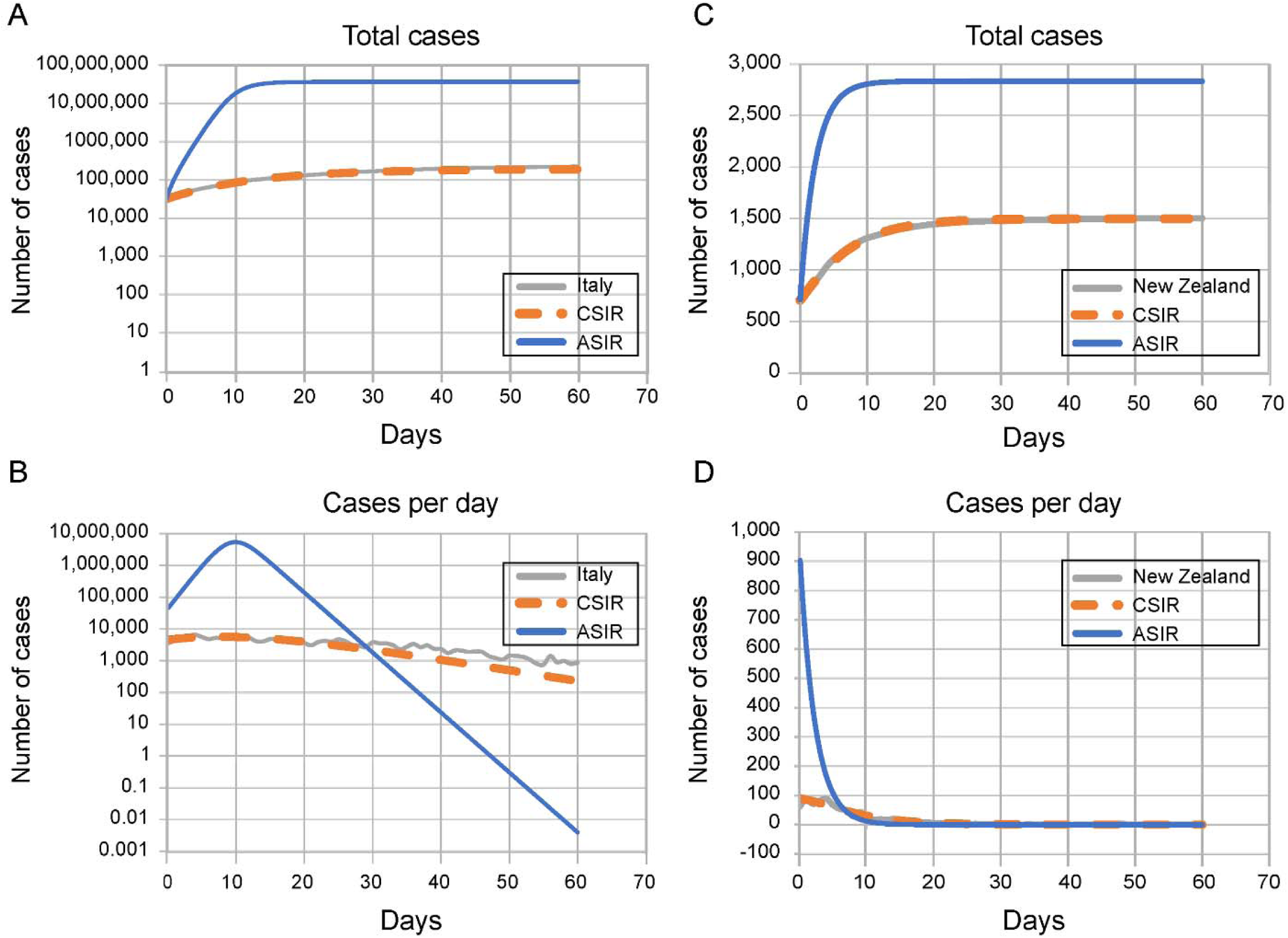
ASIR and CSIR simulations of the Italy (9A & B) and New Zealand (9C & D) data. 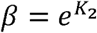 where *K*_2_ is from Table 1 and γ is equal to the *K*_*1*_ constant from Table 1.

In a second step, we recast Equations S1-49 and S1-50 in terms of *β*(*t*) and *γ*(*t*):

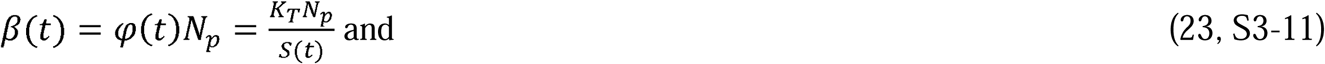

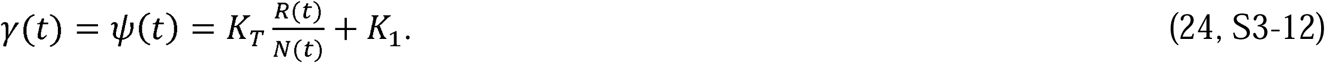

Using Equations 23 and 24, we then calculated the time series of *β* and *γ* necessary to generate the CSIR curves in Figure 9. Those time series, plotted in Figure 10, show that *β* is nearly a constant, while *γ* clearly is not.

**Figure 10.**
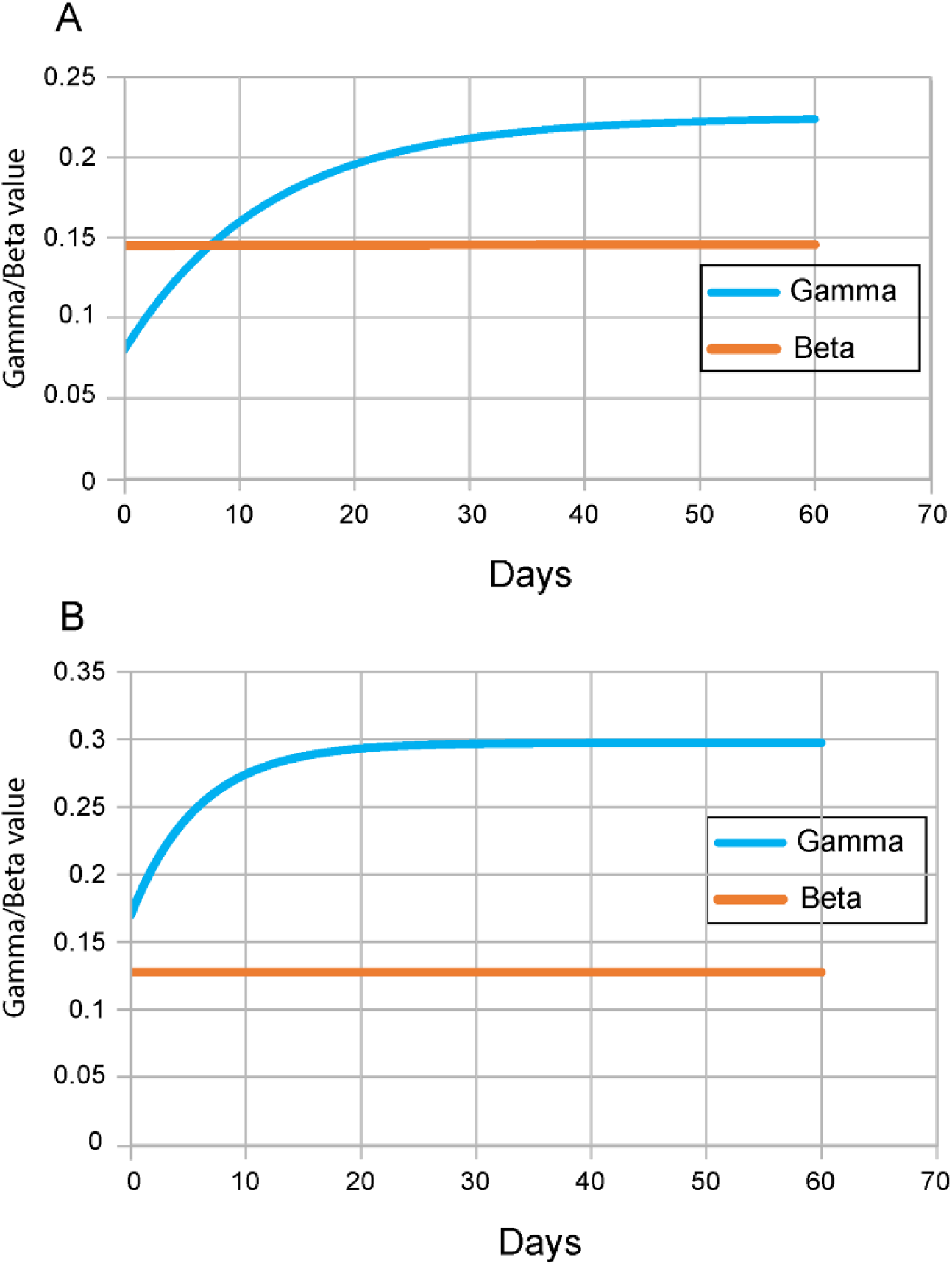
Time series for *γ* and *β*. **A)** Italy. **B)** New Zealand. These are the values of *γ* and *β* necessary for the ASIR approximation to accurately model the country data.

As a last step in the analysis, we used the values of *β* and *γ* plotted from Figure 10 in the ASIR model to generate the curves in Figure 11. This figure shows that when *β* and *γ* are forced to vary according to Equations 23 and 24, the ASIR model fits the country data quite well.

**Figure 11.**
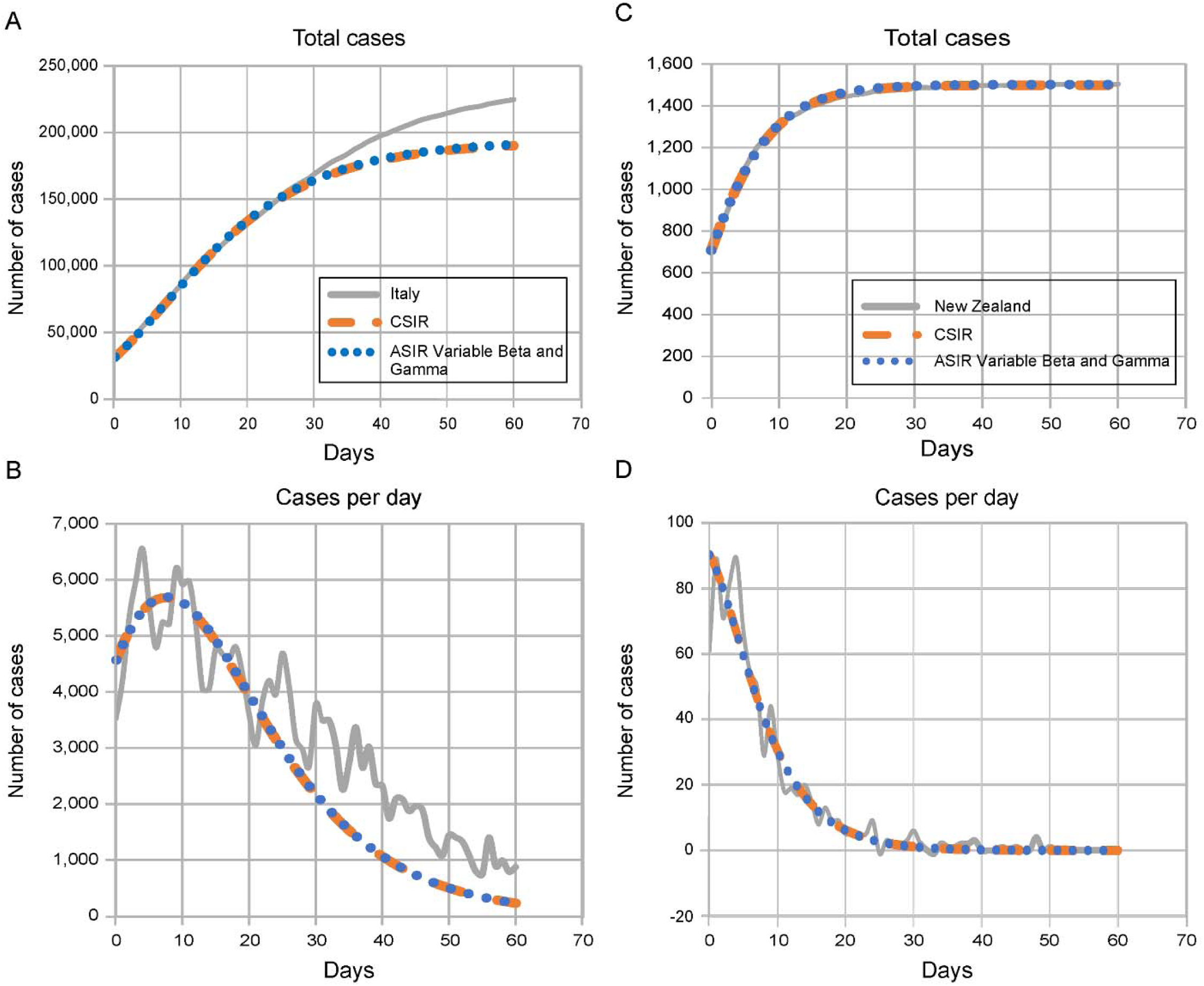
Total cases and new cases per day for Italy and New Zealand. The country data and the CSIR model plots are the same as in Figure 9. The ASIR Variable *β* and *γ* plot uses the *β* and *γ* values from Figure 10 in the ASIR equations.

Equations 23 and 24 themselves elucidate why the ASIR model, with constant *β* and *γ*, is not an accurate or appropriate approximation. Equation 23 makes it clear that when *S*(*t*)decreases by a significant percentage, neither *β* nor *φ* can be appropriately modelled as constants. Also, as can be seen in Equation 24, the assumption that *γ*, and therefore *ψ*, is a constant ignores the effect of the growing recovered (and therefore resistant) fraction, 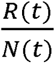, of the subpopulation, *N*(*t*), on the epidemic dynamics.

Figures 7 and 11 provide ultimate validation of the veracity of the CSIR solution. In Figure 7, we show that the CSIR solution can be configured to replicate ASIR simulations by embedding the ASIR approximations within the CSIR framework. In that setting, the two piecewise and logically invariant CSIR parameters, K_T_ and P_c_, are forced to take implausible and unrealistic time courses. Figure 11 demonstrates, in counterpoint, that an ASIR model can produce results identical to a CSIR model if the analogs of the CSIR parameters (*β* and *γ*) are permitted to vary in time according to Equations 23 and 24. Equations 23 and 24 together serve as a correction to the ASIR model.

Only the CSIR model produces results consonant with real-world data. The ASIR model fits reality only when “coached” to a time variation for its two parameters, which requires insight derived from the CSIR solution.

### 2.4 Using the CSIR solution to control epidemics

As the preceding analysis demonstrates, early and strong containment actions will end the epidemic correspondingly early, reducing the number of cases exponentially. The peak in daily cases will never be delayed by strong actions, but will, in fact, always come earlier. The curve does <u>not</u> flatten with strong intervention. It has an earlier peak and an asymmetric tail that falls to an earlier end, as shown in Figure 6D.

Fortunately, the utility of the CSIR model solution goes beyond merely predicting that the epidemic will end earlier with the imposition of strong social measures. With an exact solution to the CSIR model in hand, we can also derive important, heretofore unknown relationships and identities. These are numerous and are described in detail in Supplements 4, 5, and 6. They are supported using real life and hypothetical examples. In these supplements we develop identities that illustrate the importance of early intervention strategies and we derive mathematical tools that public health officials can use to characterize, control, and end an epidemic.

If, however, an epidemic is not controlled in its early stages, using easily obtainable data (total cases and new daily cases), the tools explicated in Supplement 6 can be used to determine the state of the epidemic and to quantitate actions that might be taken to control and end the epidemic within a desired and predetermined period of time. Strong intervention actions are likely necessary and, using the RCO metric, the effectiveness of these actions can be quickly determined from the resultant case trends. Public health officials can closely monitor the epidemic state and adjust control measures accordingly.

As an epidemic progresses, outbreaks and surges should be expected. These will occur if containment flags, more infectious variants emerge, or new cases are introduced from outside the region of focus. Each of these contingencies needs to be controlled. As explained in Supplement 6, the start of an outbreak causes obvious changes in the behavior of the RCO metric. Diligently monitored, these RCO changes can be detected early enough for public health officials to react in a timely manner to bring outbreaks under control and to re-establish the planned course.

## 3. Discussion

Data on the current COVID-19 pandemic are widely accessible from a variety of sources, updated daily. The unfolding panorama provides a test bed for models that predict outcomes and the effects of various interventions. Because different countries employed different containment strategies (3-5, 9), the world is conducting a de facto epidemiological experiment on a grand scale.

The CSIR solution to the original Kermack and McKendrick (1) integro-differential equations accurately projects the epidemic trends. It makes clear that short and sharp containment measures produce a rapid truncation of upward case trends, thereby shortening—not lengthening—the time needed to bring epidemics under control. An indicator of the rate of change in epidemic dynamics (e.g., the RCO) allows direct observation of the effectiveness of intervention measures and provides policy makers with an opportunity to react before new outbreaks gain momentum.

Current epidemiological models use the ASIR approximation, which has long been assumed to be a reasonably accurate representation of the complete equations developed in 1927. ASIR models exist in many variants, both deterministic and stochastic, and their behavior is widely known. It is startling, then, that when the ASIR model is tested using the currently available data from the COVID-19 pandemic, it fails the most basic test for any model by projecting trends that are opposite of those easily visible in the data from multiple countries. The reason for this failure is quite simple: the ASIR equations embody inherently implausible assumptions about the disease and population dynamic behavior.

Unfortunately, despite its failure to accurately model real-world trends, the ASIR model and its variants have been used to fashion guidelines for epidemic management. Tragically, the false notion that stronger containment measures lengthen the pandemic may well have caused leaders, especially those most concerned with economic performance, to see containment measures as producing only a modest reduction in the horror of an epidemic peak while significantly prolonging economic disruption. Not only do ASIR models project incorrectly that epidemics are prolonged by containment; but they significantly underestimate the reduction in the peak of cases that containment provides.

Every country and economy can use the CSIR solution presented here to plan and implement the highest level of containment measures deemed sustainable to quickly reduce case numbers to levels at which case identification, contact tracing, testing, and isolation can be maintained, allowing a more rapid return to nearly normal social activity and minimizing economic consequences. The ultimate insight from the CSIR solution is one of hope: the path of an epidemic is not an uncontrollable force of nature, nor is epidemic control inevitably the road to economic ruin. Rather, the afflicted population can, through their behavior, choose to control their destiny.

## Data Availability

The data will be made available by the authors upon request

## Abbreviations used in this text

ASIR: approximate SIR
CSIR: complete SIR
RCO: rate of change operator
RMM: residential mobility measure
SIR: susceptible–Infectious–Recovered

## Author contributions

T. Duclos and T. Reichert: Conceptualization, Writing – Original draft preparation, Writing – Review & Editing. T. Duclos: Methodology, Validation. T. Reichert: Data Investigation.

## Supplementary Material

### Supplement 1. The complete SIR model and its closed-form solution

Equations are numbered sequentially within each supplement. Equation numbers from the main text appear in bold.

#### S1.1 The complete SIR (CSIR) model

The original equations from Kermack and McKendrick (1) can be written in the following form:

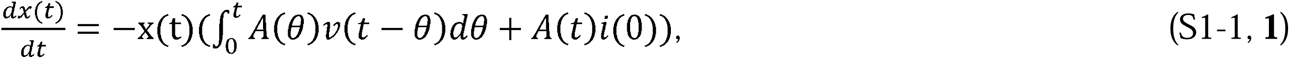

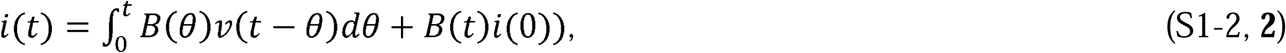

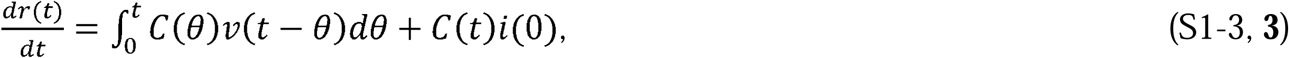

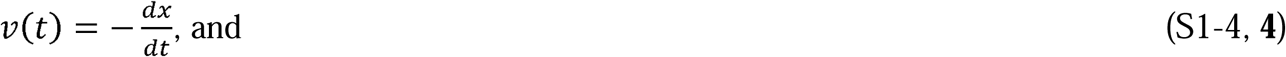

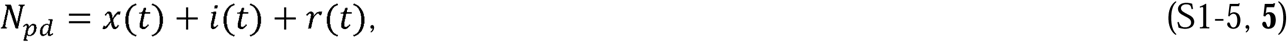

where 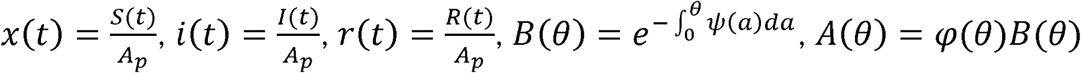, and *C*(*θ*) = *ψ*(*θ*)*B*(*θ*). Kermack and McKendrick (1, p. 703) defined *φ*(*θ*) as “the rate of infectivity at age *θ*” (page 703), and *ψ*(*θ*) as “the rate of removal” (page 703) of the infected population. We define *N*_*pd*_ as the population density; *A*_*p*_ as the area that contains the population; *S*(*t*) as the number of people susceptible to infection at time *t, I(t* as the number of people infected at time *t, R*(*t*) as the number of people recovered at time *t, N*_*p*_ as the total number of people in the population; and, therefore, *N*_*pd*_*A*_*p*_ = *N*_*p*._ The variables, θ and *a*, are dummy variables for time with *θ* defined as the stage of the infection (i.e., *θ* = the time interval since the infection).

The terms *φ*(*t*) and *ψ*(*t*) were not explicitly defined by the authors, however, they are the forms that *φ*(*θ*) and *ψ*(*θ*) take in the time domain. Excluding when *t* = *θ* = 0, the form of *φ* and *ψ* are expected to differ in the time (t) and theta (*θ*) domains.

Although Kermack and McKendrick labelled *φ*(*θ*) as “the rate of infectivity at age *θ*” (page 703) close inspection of the relationship wherein they first used this term reveals that *φ*(*θ*) cannot merely be a rate with the units of inverse theta. Rather, dimensional analysis requires that *φ*(*θ*) have the unit infectivity per contact density (contacts per unit area). Kermack and McKendrick also chose the number of susceptibles per unit area (*x*_*t*_ in their notation, *x*(*t*)in ours) as the contact density for the infected population. As a consequence, if the transmissibility of the disease does not change during the analysis, this choice for the contact density, and the definition of *φ*(*θ*) as a rate per contact density, compel *φ*(*θ*) and *φ*(*t*) to increase as time and theta grow larger because the susceptible population density perforce must decrease as the epidemic progresses. These consequences were not addressed by Kermack and McKendrick, and possibly were not recognized.

A solution to Equations S1-1 to S1-5, which correctly represents the epidemic dynamics of COVID-19 in various countries, is attainable; however, the forms of *φ*(*θ*) and *φ*(*t*) that appear in these solutions, under the constraints imposed by holding the disease transmissibility and population contact density constant, are quite extraordinary. Therefore, during the analyses in the supplements, we explicate these forms and why they arise.

#### S1.2 The solution to the CSIR model

Equations S1-1 to S1-3 can be reformulated to make a solution easier to find. The first step is to define two functions, μ(*t*) and ρ(*t*), as

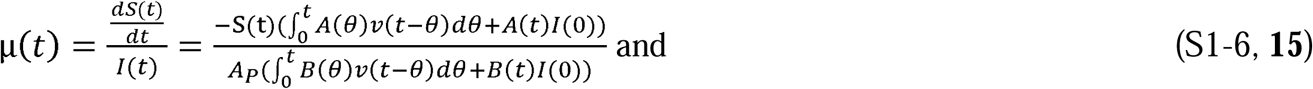

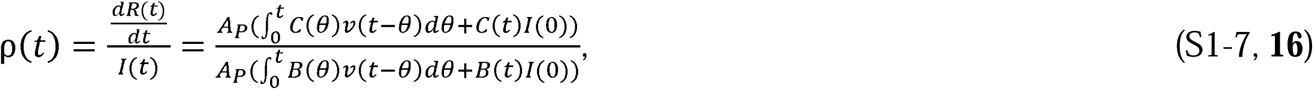

and then rewrite Equations S1-1 to S1-3 after multiplying each by *A*_*P*_:

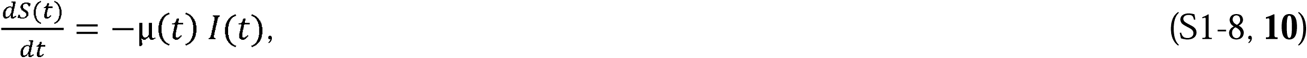

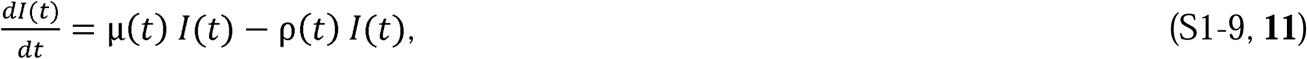

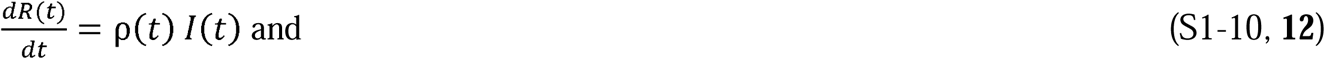

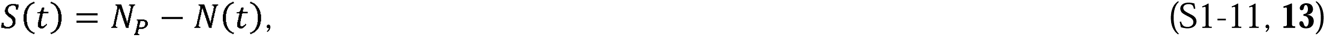

where 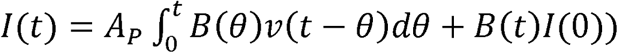, *N*(*t*) = *I*(*t*) + *R*(*t*) = the number of people that have been infected up until time, t, and 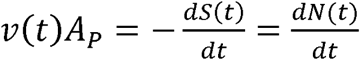.

This set of equations is mathematically equivalent to the Kermack and McKendrick (1) system of Equations, S1-1 to S1-5.

To solve Equations S1-8 to S1-11, we first find an expression for 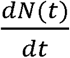 in terms of N(t), the number infected to date, the transmissibility of the disease, and the behavior of the population. We then solve this equation for *N*(*t*). Once we have determined *N*(*t*), we find expressions for *I*(*t*), *R*(*t*), *S*(*t*), μ(*t*), and ρ(*t*) and show that these expressions solve Equations S1-1 to S1-5.

At the beginning of the epidemic, we can write the following difference equation to describe the change in the number of infections during the time Δ*t*:

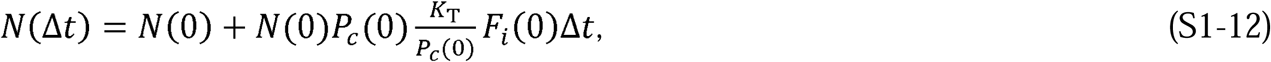

where *K*_*T*_ is the transmissibility of the disease and 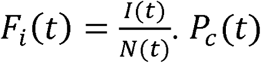 is the instantaneous number of contacts made by each member of the population *N*(*t*) and is defined by the expression 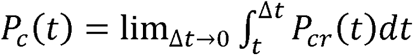, where *P*_*cr*_(*t*) is the contact rate for the subpopulation *N*(*t*).

*P*_*c*_(*t*) can be interpreted as the average number of specific infectious contacts each person within the subpopulation *N*(*t*) has with the entire population. Therefore, the number of interactions with infectious potential between the population *N*(*t*) and the entire population is *N*(*t*)*P*_*c*_(*t*), at time *t*. By “specific infectious” we mean that each person is assumed to interact only with the same people (quantity = *P*_*c*_(*t*)) in a way that might transmit the disease during the time under consideration. *P*_*c*_(*t*) is the parameter that captures the behavior of the population in the analysis going forward.

The structure of Equation S1-12 captures the dynamics of the epidemic as it begins. The quantity *N*(0)*P*_*c*_(0) is the number of potentially infectious contacts between people, *F*_*i*_(0) is the fraction of those contacts that involve already infected persons, and 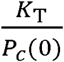 is the number of successful transmissions of the disease per infectious contact.

We also note that, using Kermack and McKendrick’s (1) formulation of the problem of epidemic origination, Equation S1-12 could have been written as

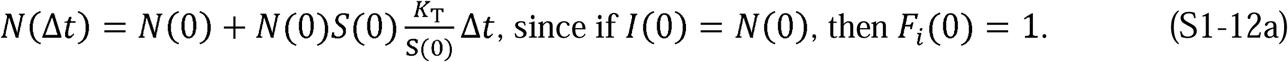

Since S1-12 and S1-12a are obviously mathematically equivalent, we ostensibly begin, mathematically, from the same place as Kermack and McKendrick. However, rather than forcing the transmissibility per contact density, *φ*, to be a relationship describing disease transmissibility per each susceptible person, we have instead defined the function describing transmission, 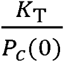, as the transmissibility per the actual number of contacts by the population N(t). This subtle reformulation allows us to develop both a solution and many useful relationships.

It is reasonable to assume that the transmissibility of the disease, *K*_*T*_, will remain constant for an interval of time because it depends only on properties of the agent causing the disease. Adopting this assumption, Equation S1-12 can then be rearranged as

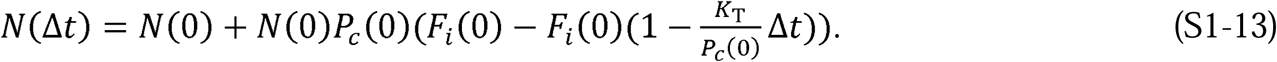

In Equation S1-13, since *N*(0) is a given and *P*_*c*_(0) is only a function of people’s behavior, the quantity 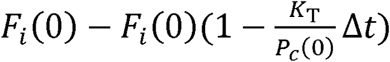 must be the change in *F*_*i*_(0) during the time period Δ*t*. That is, the difference between *F*_*i*_(0) and *F*_*i*_(Δ*t*) and can be written as

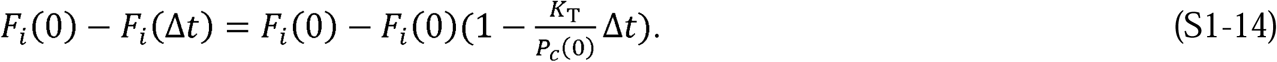

This can then be rewritten as

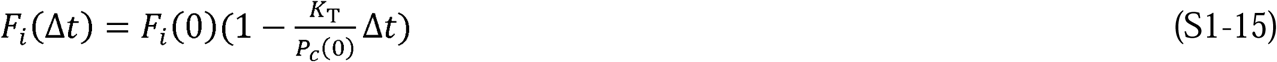

and for any time (n+1)Δ*t*, S1-15 can be written as

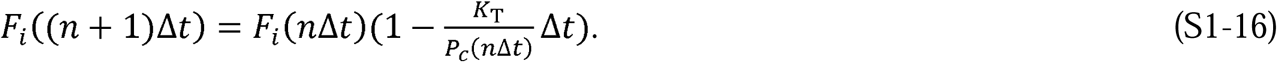

Using S1-16, we can write the following for *N*(2Δ*t*):

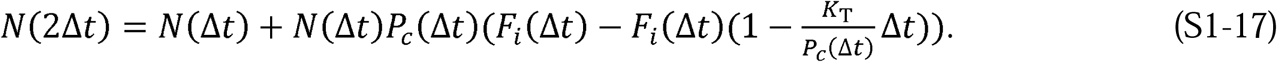

This can be rewritten as:

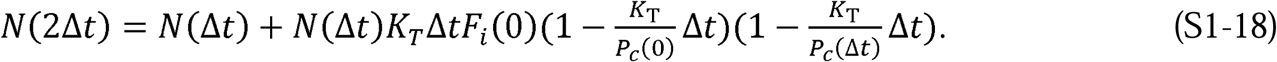

This process of substitution can be repeated to develop the following relationship:

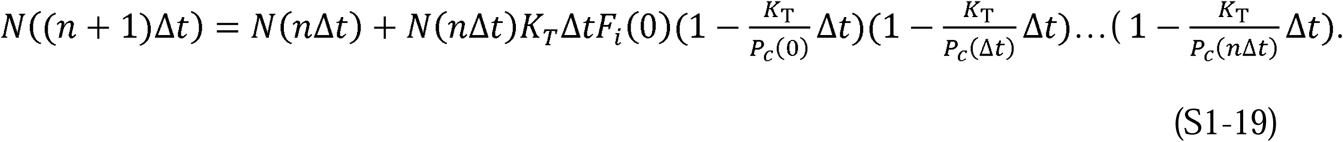

Defining *n*Δ*t* = *t* and allowing Δ*t* → 0, Equation S1-19 becomes

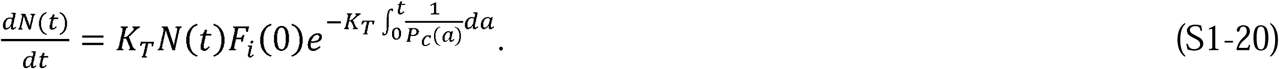

Equation S1-20 is easily solved for *N*(*t*):

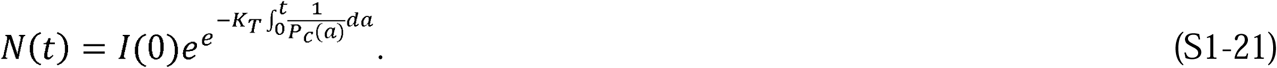

This provides us with a solution to Equation S1-8:

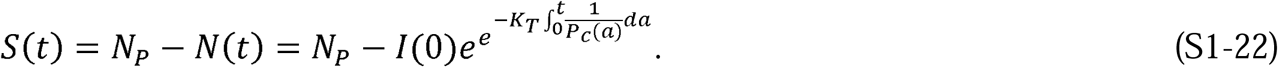

To completely solve the system of equations in Equations S1-8 to S1-11 we now need to find expressions for *I*(*t*), *R*(*t*), μ(*t*), and ρ(*t*). We begin by first noting that

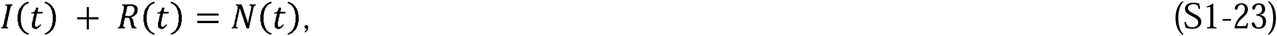

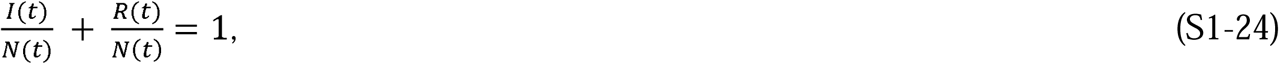

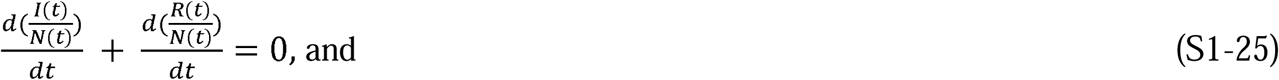

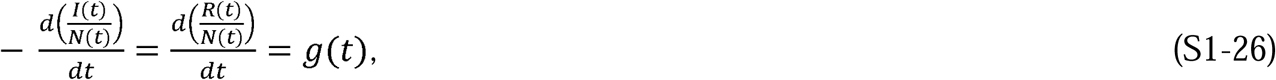

where *g*(*t*) is a yet unknown function of time.

Since all the recovered people must have been infected, 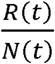 can only change if *I*(*t*) changes, and only to exactly the extent that the quantity 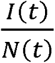 changes. Thus, we can rewrite S1-26 in the following form:

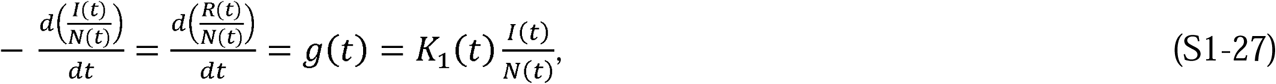

where *K*_1_(*t*) is a function that modifies the fraction 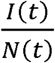 in association with recoveries at time *t*. It is initially assumed to be a function of time.

Equation S1-27 is a simple differential equation in the variable 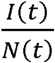, whose solution is

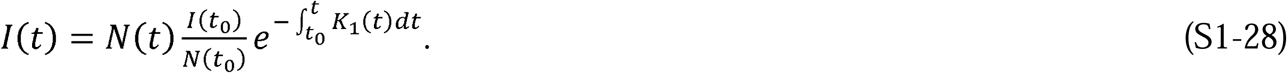

Or, if *K*_1_(*t*) is a constant,

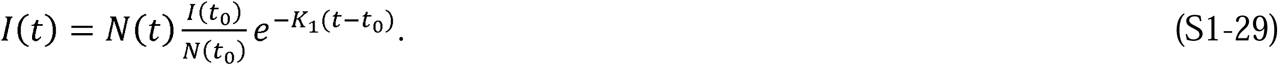

Likewise, the solutions for *R*(*t*) are

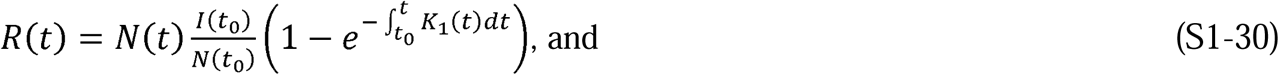

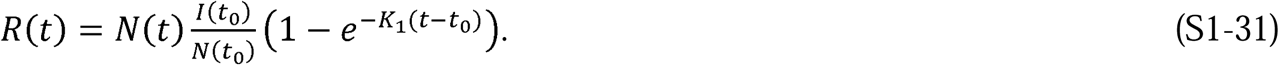

When time *t*_0_ = 0, then *N*(*t*_0_) = *I*(*t*_0_) = 1 and Equations S1-28 to S1-31 become

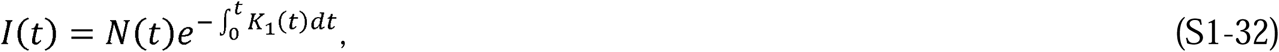

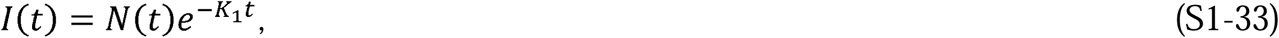

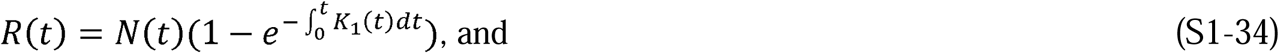

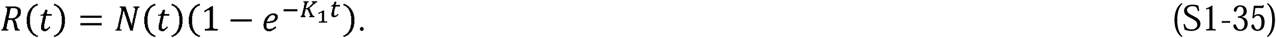

We must now define *K*_1_(*t*) in terms of the epidemic parameters *P*_*c*_(*t*) and *K*_*T*_. By forcing Δ*t* to go to zero in Equation S1-15 we arrive at the following differential equation:

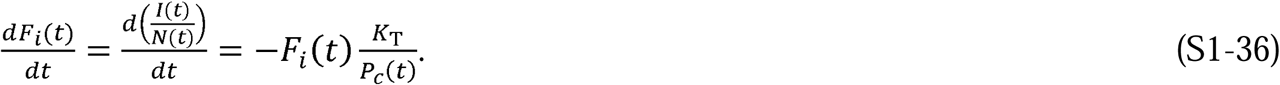

Comparing Equations S1-27 and S1-36, we can see that:

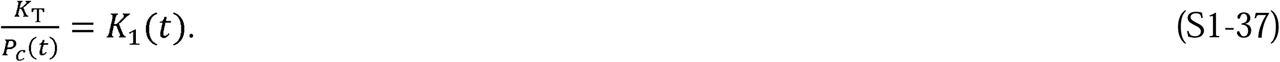

Since the population’s behavior is likely to remain constant for many days running, for the initial model development, we assume that *P*_*cr*_(*t*) is constant and, therefore, that *P*_*c*_(*t*) and *K*_1_(*t*) are also constants. Of course, we only expect this to be true on a piecewise basis because eventually, in response to the epidemic, populations adopt measures such as mask wearing, online meetings, quarantines, and reopening. These measures significantly change the contact rate, and therefore *P*_*c*_(*t*). The consequences of piecewise variation of *K*_1_(*t*) are explored and explained in Supplement 6.

Having defined *K*_1_(*t*) as a piecewise constant, for simplicity, we refer to it as *K*_1_, and to *P*_*c*_(*t*) as *P*_*c*_ for the remainder of this portion of the analysis. We can now find the solution to Equations S1-8 to S1-10 and S1-1 to S1-3.

Using the assumption that *K*_1_(*t*) and *P*_*c*_(*t*) are constants, the expression for 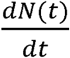 from Equation S1-20, and Equation S1-37, we arrive at the following:

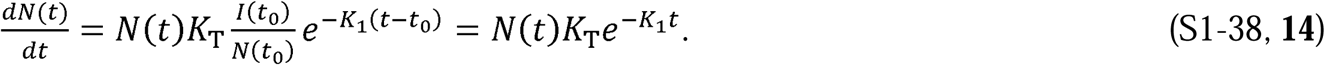

Integrating S1-38 produces the expression for the total number of infections, *N*(*t*):

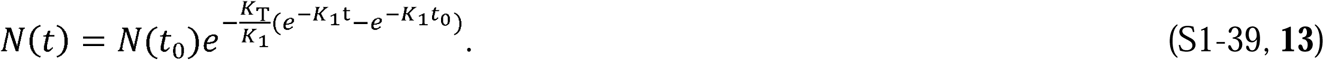

We can now write expressions for *I*(*t*), *R*(*t*), and *S*(*t*) as

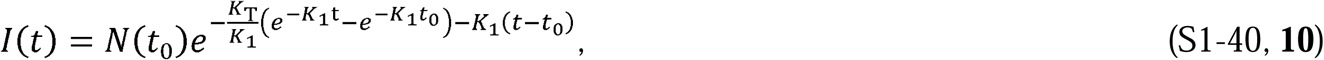

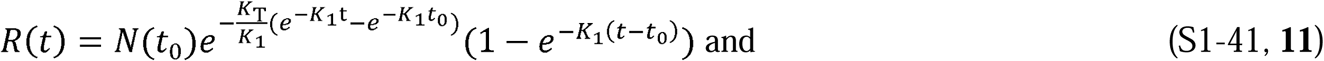

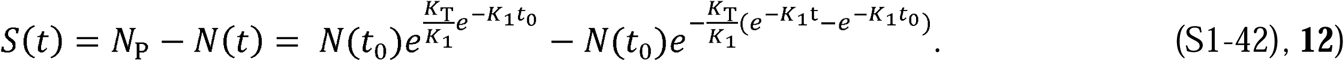

Equations S1-40 to S1-42 can each be differentiated and rearranged to produce these differential equations:

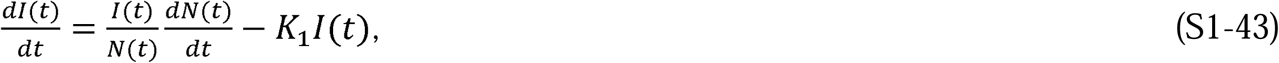

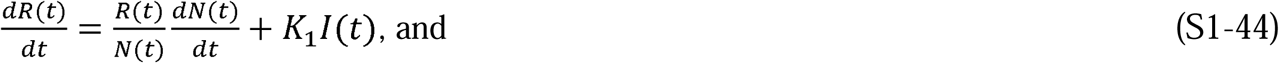

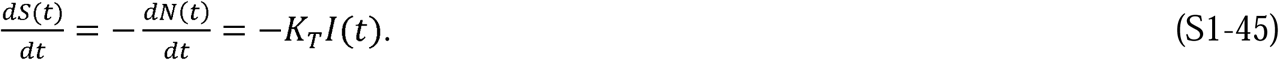

Comparing Equation S1-45 to Equation S1-8, we conclude that *K*_T_ = μ(*t*). If we also equate Equations S1-44 and S1-10, we can find an expression for *ρ*(*t*):

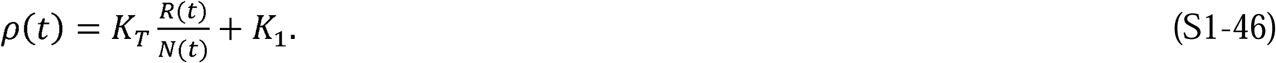

Using these definitions for μ(*t*) and ρ(*t*), we can see that Equations S1-43 to S1-45 are equivalent to Equations S1-8 to S1-10 and, by extension, to Equations S1-1 to S1-3. Therefore, since S1-40 to S1-42 solve Equations S1-43 to S1-45, they are also solutions to Equations S1-8 to S1-10 and, again by extension, solutions to Kermack and McKendrick’s (1) original Equations, S1-1 to S1-3.

#### S1.3 Proof that the proposed solution solves the original Kermack and McKendrick integro-differential equations

As an affirmative demonstration that the expressions in Equations S1-40 to S1-42 solve the original Kermack and McKendrick equations (1), we now insert these expressions into the original equations and show that they are satisfied. To accomplish this demonstration, we must first determine the form of the expressions for *φ*(*t*), *ψ*(*t*), *φ*(*θ*), and *ψ*(*θ*).

Using Kermack and McKendrick’s formulation, *φ*(*t*) and *ψ*(*t*) can be defined as

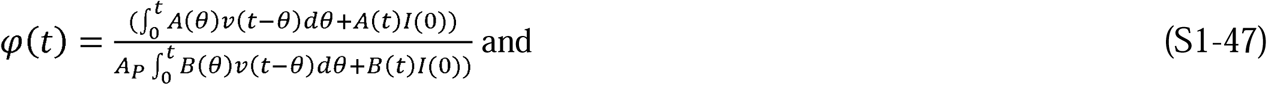

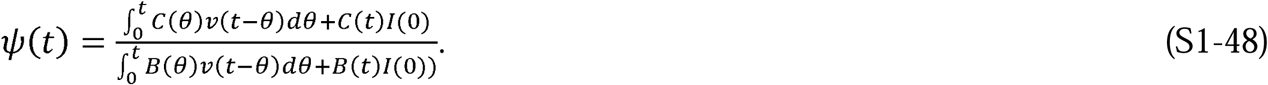

From these definitions and the prior analysis, we can clearly see that

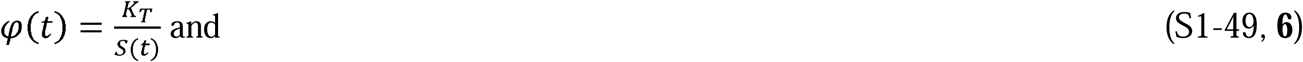

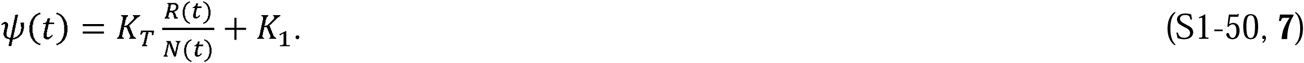

We now relate *v*(*t* −*θ*) to the epidemic parameters and our adopted naming conventions. In Kermack and McKendrick’s formulation, *A*_*p*_*v*(*t* −*θ*), is the number of infections new at time t. In our formulation, this is 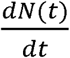. Therefore,

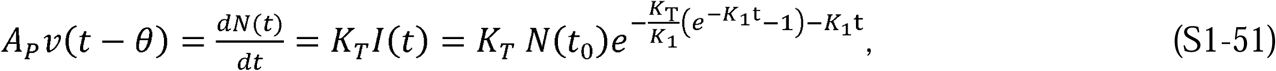

which adopts Kermack and McKendrick’s convention that t(0) = 0.

Since infections enter the infected population at the rate *K*_*T*_ (see Equation S1-45), it is reasonable to choose *K*_*T*_ as the rate at which infections exit the infected population in theta space. Therefore, we choose the following expression for *ψ*(*θ*):

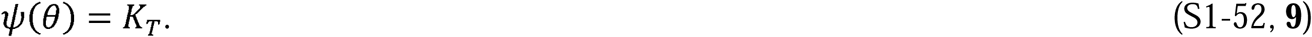

From S1-52, we can calculate *B*(*θ*):

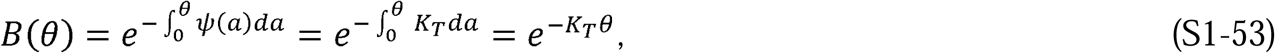

where *a* is a dummy variable.

Equation S1-50 can be rewritten in terms of the epidemic parameters as

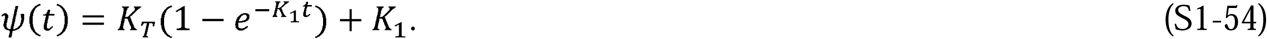

This allows us to calculate *B*(*t*):

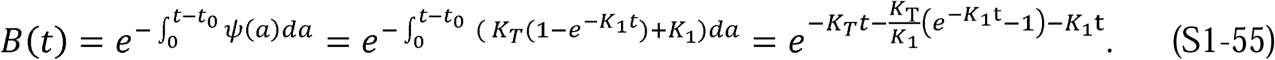

We can now solve for I(t) using Equations S1-2, S1-51, S1-53, and S1-55 after multiplying both sides of S1-2 by *A*_*P*_:

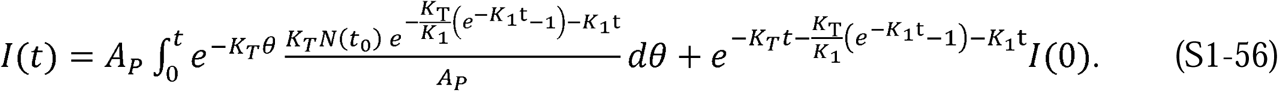

Since the term 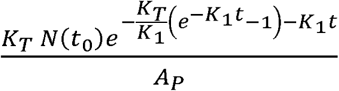 is not a function of *θ*, it can be brought outside the integral and Equation S1-56 becomes

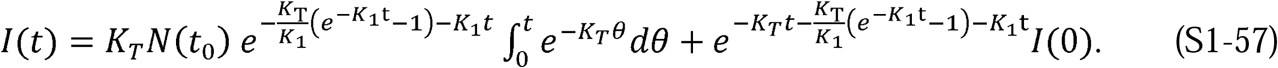

Since *I*_0_ = *N*(*t*_0_), Equation S1-57 is easily solved to find the following expression:

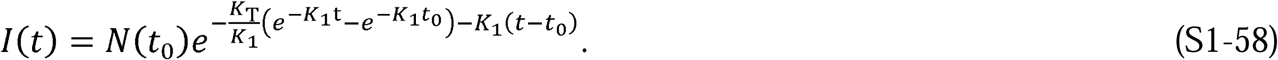

In a similar fashion, we can use Equations S1-3, S1-51, S1-53, and S1-55 to find the following:

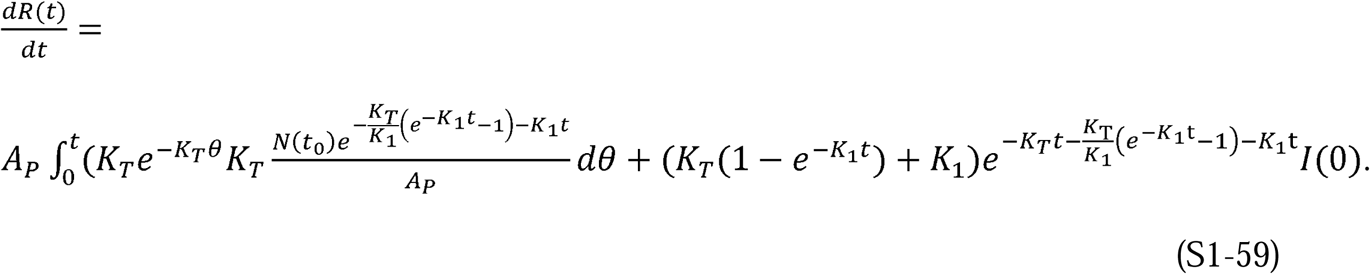

Using the same approach as in the solution for *I*(*t*), we can solve Equation S1-59 to find the expression for 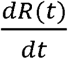:

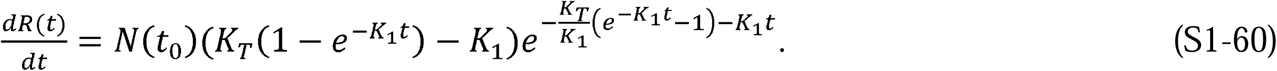

Equation S1-60 can be integrated to find the expression for *R*(*t*):

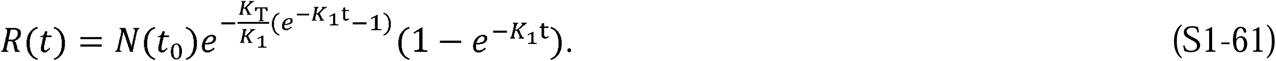

To solve Equation S1-1, we must find the form of *φ*(*θ*). Although, finding the form of *φ*(*θ*) is not as straightforward as was the route to the form of *ψ*(*θ*), we can make a few a priori statements regarding our expectations of its form. First, we expect *φ*(*θ*) to increase with increasing theta, just as *φ*(*t*) increases with time. Second, we expect the increase with theta to be larger than the increase of *φ*(*t*) with time since the *v*_*t,t*_ (using Kermack and McKendrick’s notation of *v*_*t,θ*_) portion of the infected population, *v*_*t,θ*_, is a subpopulation that we expect to decrease at a rate faster than that of the overall population. This subpopulation will have been infected the longest and, therefore, recovers at a high, possibly maximal rate.

With these expectations in mind, we write the following using Equations S1-1, S1-45, and S1-51:

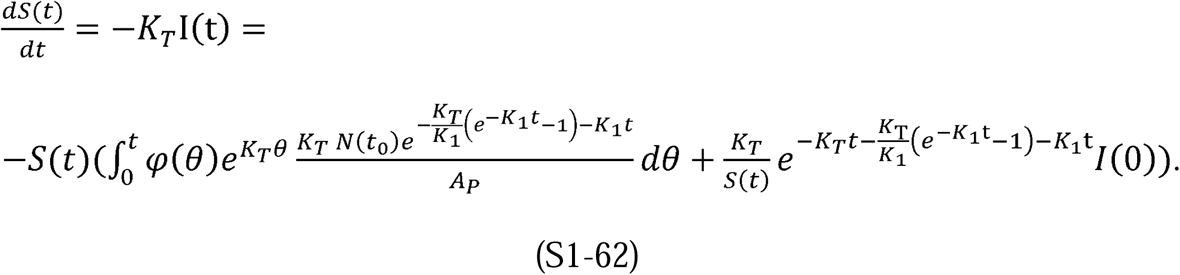

This can be simplified to

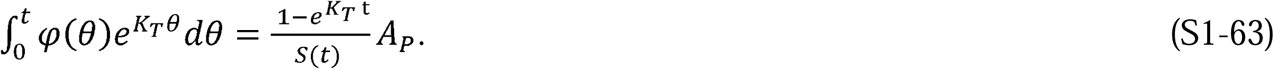

We can then easily show that the following expression for *φ*(*θ*) satisfies S1-63:

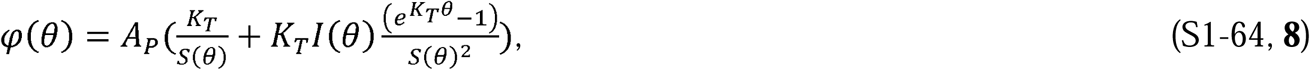

where 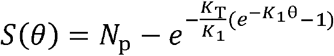 and 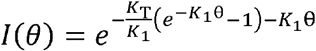.

As expected, *φ*(*θ*) grows with increasing theta in Equation S1-64 and it grows at a faster rate as theta increases than *φ*(*t*) grows with time in Equation S1-49.

Now we can use Equations S1-1, S1-51, S1-55, and S1-65 in the same fashion as before to find the expression for 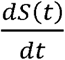:

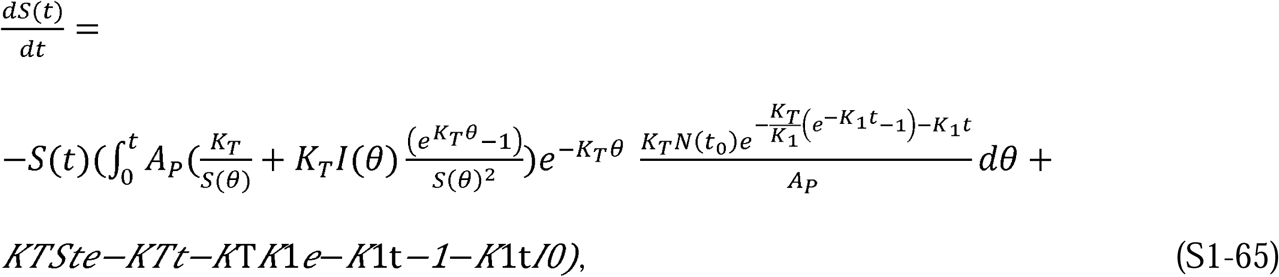

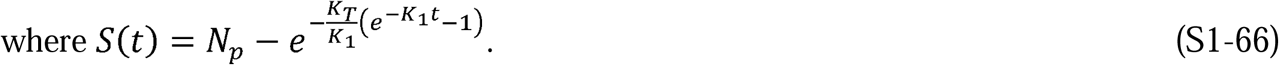

Taking the functions dependent on *t* outside the integral, simplifying and solving the integral we obtain the following:

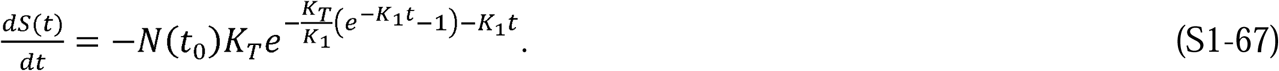

Equation S1-67 can then be integrated to obtain the expression for S(t):

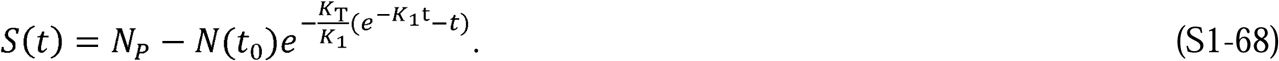

The expressions for I(t), R(t), and S(t) in Equations S1-58, S1-61, and S1-68 are the same as those in S1-40 to S1-42. Therefore, Equations S1-40 to S1-42 are a solution to the original Kermack and McKendrick (1) equations.

### Supplement 2. Verification of the CSIR model

Equations S1-38 and S1-39 can be combined purposefully to define an important relationship:

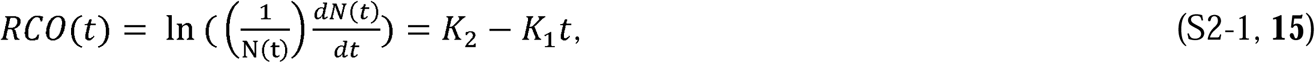

where 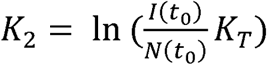 and *t*_0_ is the time between the start of the epidemic and the time at which the first data point in the epidemic is measured.

The RCO expression is convenient in that it transforms Equation S1-38 into an equation linear in *K*_1_. We call the expression the rate of change operator because it is the rate of change of new cases scaled by the current total of cases. It is Equation S2-1 that we fit to the nine data points after the imposition of containment measures in the country data in Figure 1. We also used this equation to find the parameters in Table 1.

We developed the solutions to Equations S1-1 through S1-5 assuming that *K*_*T*_ is a constant dependent only on the disease and is the same for all countries. We also assumed that *P*_*c*_(*t*) is dependent on the population’s behavior. We used these assumptions when generating the correlations of the case data for various countries in Figures 2 and 3. Both of these assumptions can be tested independently of the country correlations.

To demonstrate that *K*_*T*_ is indeed a constant, we need to first further define *P*_*c*_(*t*). As previously defined, *P*_*c*_(*t*) is the number of specific infectious contacts a member of subpopulation *N*(t) has across the entire population. This is a function of the population’s behavior. Initially, we assume this is a function of population density and further, that people’s mobility extends over a constant average effective area per unit of time. We define this area as the effective area rate, *A*_1*r*_(*t*). Using these definitions, we can write an expression for *P*_*cr*_(*t*):

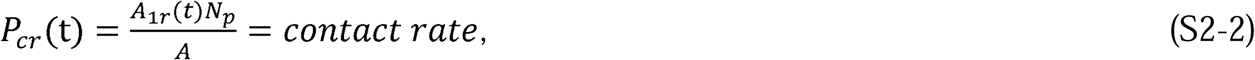

where *N*_*p*_ = the entire population of the region with the infection, *A* = the area of the region, and 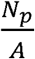 = the population density. From Equation S2-2, we can see that *P*_*cr*_(*t*) is proportional to both the population’s behavior, *A*_1*r*_(*t*), and the population density, 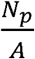.

Similar to the way we defined *P*_*c*_(*t*) using *P*_*cr*_(*t*), we now define a quantity, *A*_1_(*t*), in terms of *A*_1*r*_(*t*):

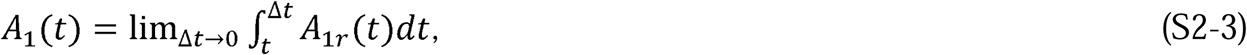

where *A*_1_(*t*)is the effective specific area traversed by an individual. In this case, “specific” has the same meaning as it has for *P*(*t*)_*c*_; that is, each person traverses only and exactly the same area for the duration of the time under consideration. We also call *A*_1_(*t*) the “effective area” because the population is typically only dispersed within ∼1% of the land within a given region of a country (12). If we take this into account, then 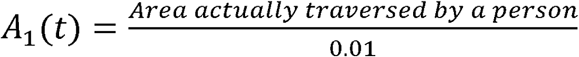.

From the preceding discussion, we can now write an expression for *K*_1_(*t*):

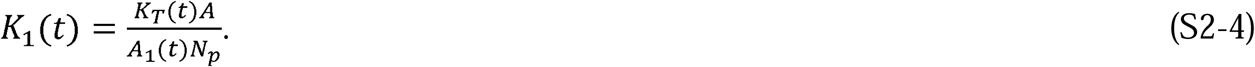

The physical meanings of *P*_*c*_(*t*) and *A*_1_(*t*) may not be intuitive. As defined, neither are rates, but they can both vary in time and their values depend on the population’s behavior. They are, respectively, the number of specific infectious people who have been contacted and the specific area traversed by any index person within a given time interval. *P*_*c*_(*t*) and *A*_1_(*t*) are constant when the number of specific infectious people or the traversed area remain constant. However, if different people are contacted within a given time interval, the rates they depend on change, and therefore, *P*_*c*_(*t*) and *A*_1_(*t*) may change even if the total number of people contacted or area covered did not change during that time interval.

We can now check the assumption that *K*_*T*_ is a constant by substituting Equation S2-4 into Equation S1-39 and solving for *K*_*T*_t. Doing this, we find the following expression:

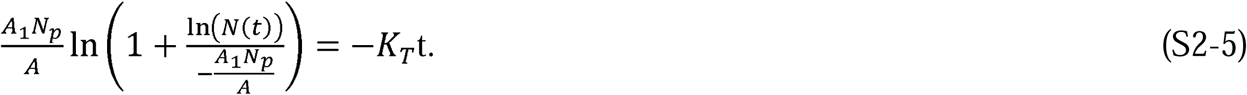

If we define 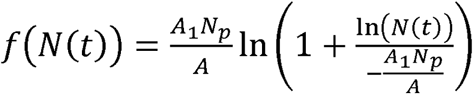, then we can also write this expression as

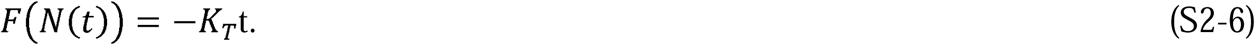

If *K*_*T*_ is a constant, then Equation S2-6 predicts that *F*(*N*(*t*)) is a linear function of time. Excepting *A*_1_, all the quantities on the left-hand side of Equation S2-5 can be found for each country in the time before containment measures are enacted and are listed in Table 2. Therefore, we can estimate whether *K*_*T*_ is a constant by first using Equation S2-5 (or S2-6) to find if there is a constant value of *A*_1_ that creates a straight line. An implicit assumption in this process is that the behavior of the population, *P*_*c*_, is constant and therefore *A*_1_ is constant, during the initial phase of the epidemic before containment measures were put in place.

Applying these assumptions, through a process of iteration, a value was found for *A*_1_ (= 0.48 km^2^) which created a straight line with a correlation coefficient of 0.96 (Figure 4). Using Equation S2-5 (or S2-6) we can then determine that the slope of the line in Figure 4 indicates that the value of *K*_*T*_ is 0.26. This analysis strongly supports the assumption that is a constant.

Independent evidence that *P*_*c*_ is a measure of the population behavior can be developed by first using Equations S2-1 and S3-8 (Supplement 3) to predict that the RCO measure will be proportional to 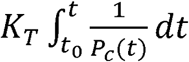. We reasoned that if an independent measure of people’s mobility during the epidemic could be found and was linearly related to the RCO, we could have additional confidence in the veracity of the CSIR solution.

Google has compiled different measures, derived from mobile phone data, of people’s mobility (11). One of these measures is termed the Residential Mobility Measure (RMM). The RMM is a measure of the percentage change in the degree to which people stayed in their residence during the pandemic relative to a baseline measured over 5 weeks starting on January 3, 2020. Since 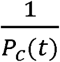 and the RMM are both inversely proportional to the population’s mobility, we hypothesized that the RMM would be a good proxy for the value of 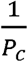. To test this, we plotted the integral over time of the daily RMM for the six countries whose data we analyzed, against the daily RCO. These plots appear in Figure 5, which clearly validates the hypothesized linear relationship.

### Supplement 3. An analysis of the ASIR model

In this supplement, we address the questions: 1) What are the implied assumptions within the conventional approximation to Kermack and McKendrick’s original equations (1), known as the SIR model? and 2) How do *β* and *γ* need to vary with time for the ASIR model equations to mimic the CSIR model?

The SIR model, with non-time-varying parameters *β* and *γ*, is described by the following equations using the same definitions of S, I, and R as in Supplement 1:

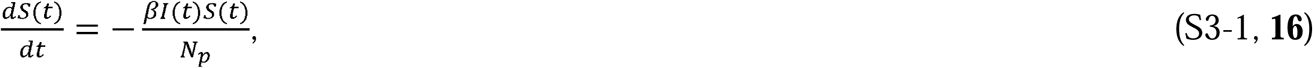

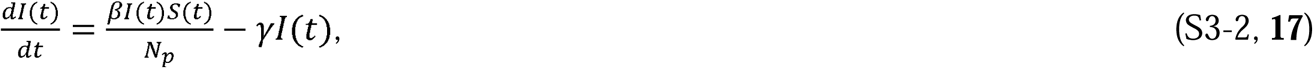

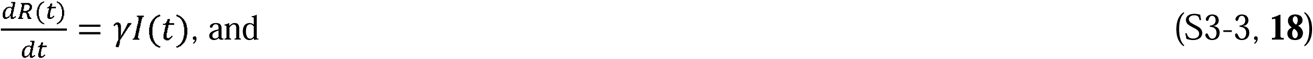

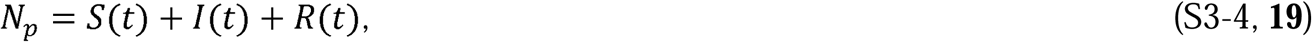

where *N*_*p*_ = total number of people in the population, *β* = rate of contact and transmission, *t*_*r*_ = time of infectiousness, and *γ* = rate of recoveries = 1/*t*_*r*_.

This formulation from Kermack and McKendrick (1) is widely promoted as illustrative of the qualitative behavior of the more complex model with time-varying parameters presented in the paper. Because the assumption that *β* and *γ* are constant strongly limits the capability to describe epidemics, we call this form of the model the Approximate SIR (ASIR) model. While not intended to model epidemic dynamics perfectly, Equations S3-1 to S3-4 have always been assumed to reflect the general trends of an epidemic (1).

To understand the implications of the ASIR approximation, we must first understand the implications of the assumption that *φ*(*t*) and *ψ*(*t*) are constant over time. The implications are easily illustrated using Equations S1-49 and S1-50. With these equations and the prior definitions that 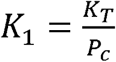 and 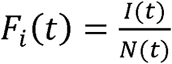, we can find expressions for the time varying *K*_*T*_(*t*) and *P*_*c*_(*t*) when *φ*(*t*) and *ψ*(*t*) are assumed to be the constants *φ* and *ψ*:

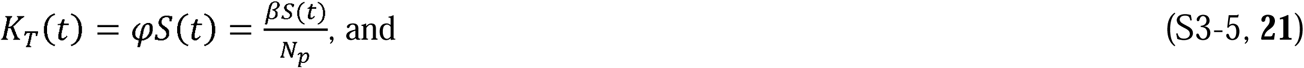

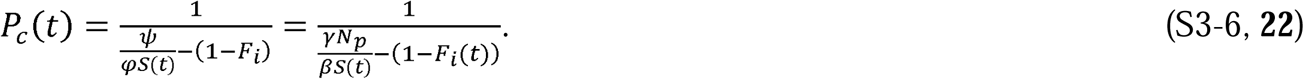

These two equations answer the first question posed at the beginning of this section. The implied assumption in the ASIR model formulation is that *K*_*T*_ and *P*_*c*_ vary with time in the manner described by Equations S3-5 and S3-6.

To apply the conditions in S3-5 and S3-6 to the CSIR solution, we must first rederive the CSIR solution under the assumption that both *K*_*T*_(*t*) and *P*_*c*_(*t*) do vary with time. We do this following the procedure used in Supplement 1, which results in the following set of relationships for *N*(*t*), 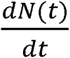, *I*(*t*) and *R*(*t*):

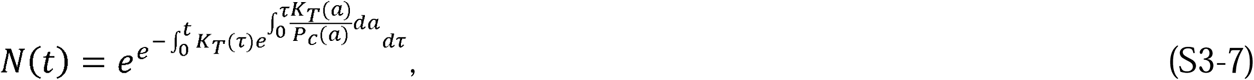

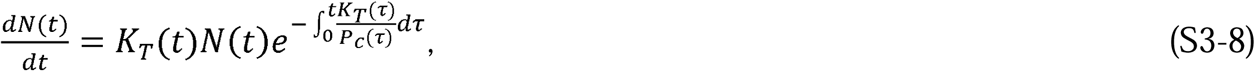

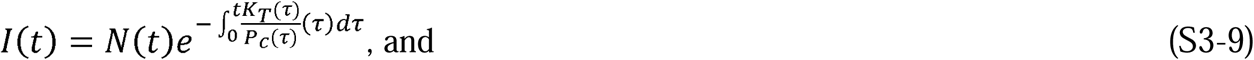

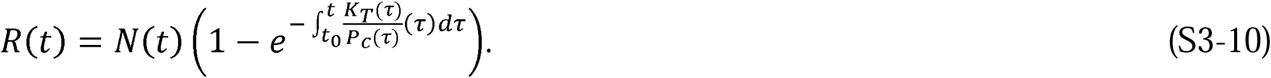

The results of the simulation of both *N*(*t*) and 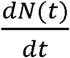 are shown in Figure 7. We created these plots by imposing the functional relationships in Equations S3-5 and S3-6, as well as using Equations S3-7 and S3-8. In Figure 8, we then used Equations S3-5 and S3-6 to plot the time variation of both *K*_*T*_ and *P*_*c*_ resulting from the assumption that *φ* and ψ are the constants *β* and *γ*, respectively.

We now explore the opposite construct, the time varying nature of *φ*(*t*) and *ψ*(*t*) (and therefore *β* and *γ*) required when *K*_*T*_ and *P*_*c*_ are constants in Equations S1-49 and S1-50. Similar to the preceding analysis, we used simulations of the CSIR solution, Equations S1-49 and S1-50, and the ASIR model. Since the CSIR solution predicts the country data well, we also compared the simulation results to the data from two countries (Italy and New Zealand) and demonstrate that only if *φ*(*t*) and *ψ*(*t*) vary as prescribed in Equations S1-49 and S1-50 can the ASIR approximation model the country data with any degree of accuracy.

We first used the values of *K*_1_ and *K*_2_ in Table 1 to derive values of *K*_*T*_ and *K*_1_ for Italy and New Zealand and used the CSIR solution to project the results. We then simulated the ASIR model with the assumption that the values of *φ* and *ψ* (and therefore *β* and *γ*) were constant and directly related to the values of *K*_*T*_ and *K*_1_ used in the CSIR solution. The results of the CSIR simulation and the ASIR simulation are plotted in Figure 9, along with the country data. The CSIR model accurately models the country data and the ASIR model does not.

In a second step, we recast Equations S1-49 and S1-50 in terms of *β*(*t*) and *γ*(*t*):

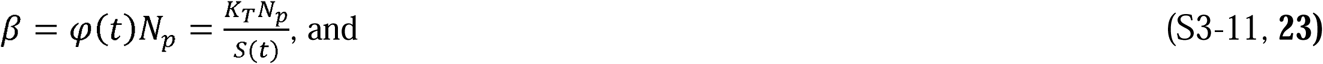

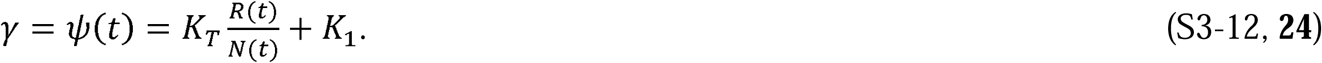

Using Equations S3-11 and S3-12, we then calculated the time series of *β* and *γ* necessary to generate the curves in Figure 9 and plotted them in Figure 10. Next, we used the values of *β* and *γ* plotted in Figure 10 in the ASIR model to generate the curves in Figure 11. Clearly, when *β* and *γ* are allowed vary in accord with the time variations of *φ*(*t*) and *ψ*(*t*), exposited above, the ASIR equations model country data quite well.

Equations S3-11 and S3-12 answer the second question posed at the beginning of this supplement. If *β* and *γ* are forced to vary according to these equations, then the ASIR model will model the epidemic. In a sense, Equations S3-11 and S3-12 are corrections to the ASIR model.

### Supplement 4. Important relationships derived from the CSIR solution

Taking the limit of Equation S1-39 as *t* → ∞, we obtain the expression for the total number of individuals who will be infected throughout the entire epidemic:

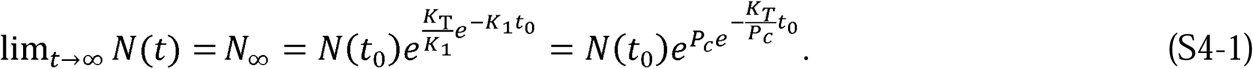

Mathematically, Equation S4-1 demonstrates that the total number of infections expected, *N*_∞_, and the behavior of the population, *P*_*c*_, are interrelated in the sense that changes in *P*_*c*_ have an exponential effect on the final number of infections. The exponential nature of the relationship underscores that small changes in population behavior dramatically affect the epidemic’s outcome. However, the reciprocity also means that the eventual number of cases produced by the epidemic is not foreordained, but rather, a strong function of interventions introduced.

We gain insight into the meaning of the CSIR model solution by looking at Equation S1-43 in more detail. Equations S1-38 and S1-40 can be used to rewrite Equation S1-43 as

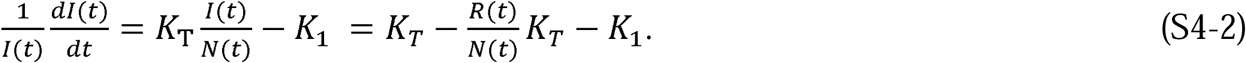

The left-hand side of Equation S4-2 is the rate of change in the number of new infections per person currently infected. The first two terms on the furthest right-hand side of Equation S4-2, 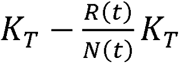, describe the net rate of successful infection. Since *K*_*T*_ is the rate at which an infected person causes infections per infectable contact, the terms 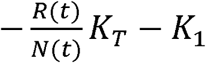 must represent the rate of recovery per infected person.

We gain an additional intuitive insight about the solution from the following relationship, derived from Equations S1-38, S1-39, and S4-1:

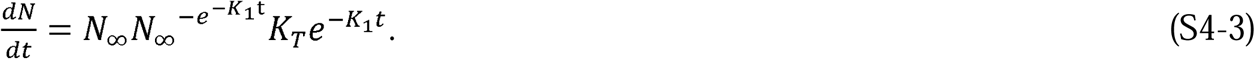

In words, the form of Equation S4-3 is

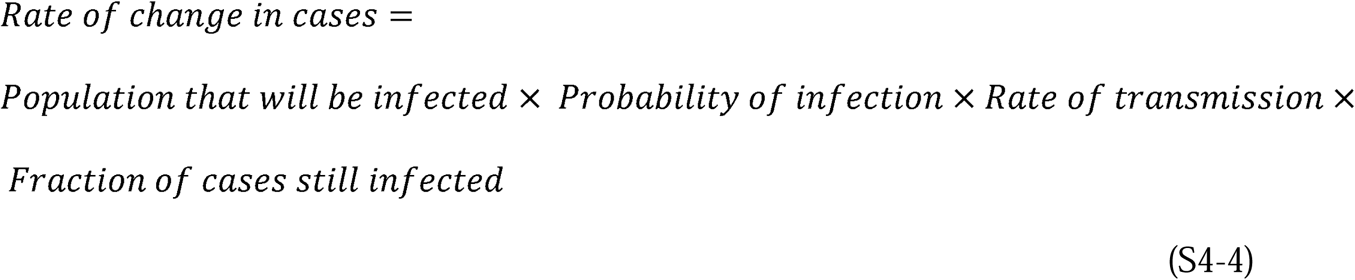

or

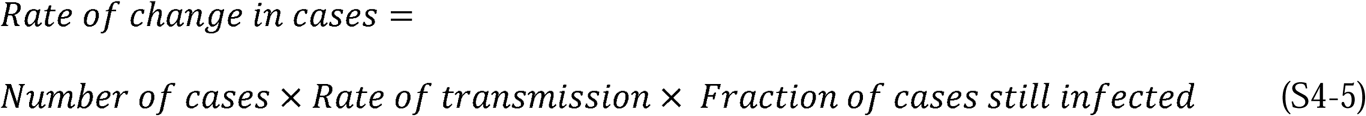

Equations S4-4 and S4-5 illustrate the logic of the CSIR solution in terms of probabilities.

Finally, using the prior definition, 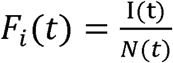, and if *t*_0_ = 0, we can use Equation S4-1 to write this simple expression for the CSIR solution for total cases:

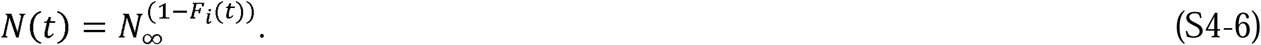

#### S4.1 Additional properties of the CSIR solution

Substituting the values for *R*(*t*) and *N*(*t*) from Equations S1-39 and S1-41 into Equation S1-46, we can arrive at an expression for ρ(*t*) as a function of time:

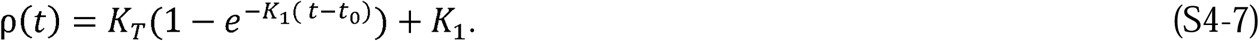

From Equation S1-9, we can see that the number of infections, *I*(*t*), will begin to decrease when 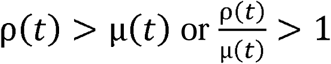.. In this way, the ratio, 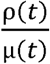, serves as the inverse of what is known in the conventional epidemiology literature as the Replication Number. We call the inverse of this ratio, the Effective Replication Number.

Using the previously developed expressions for ρ(*t*) and µ(*t*), we can write the following criteria for when the epidemic will begin to decline:

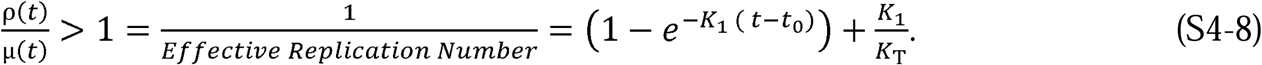

Using Equation S4-8, we obtain the following expression for when the decline begins:

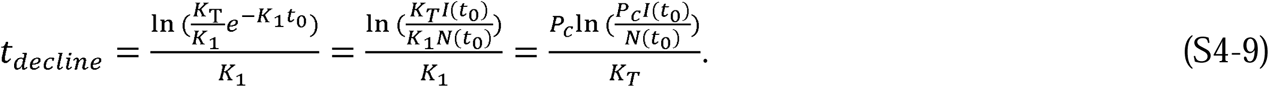

If we differentiate both sides of Equation S1-38, we obtain an expression identical to Equation S4-9:

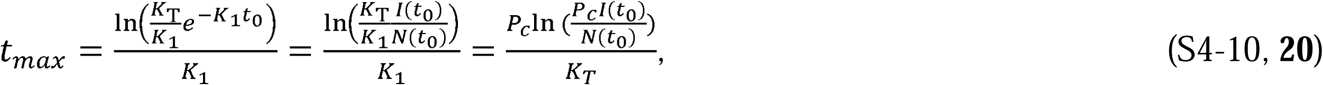

where *t*_*max*_ = the time to the peak of new infections. The time of the peak in new cases coincides, as it should, with the start of the decline of infections.

Equation S4-10 demonstrates the relationship between the strength of social intervention measures, *P*_*c*_, and the time to the peak of new infections. When social interventions are stronger (smaller *P*_*c*_), the time to the peak will always be shorter.

Another important expression is the rate of acceleration of the epidemic:

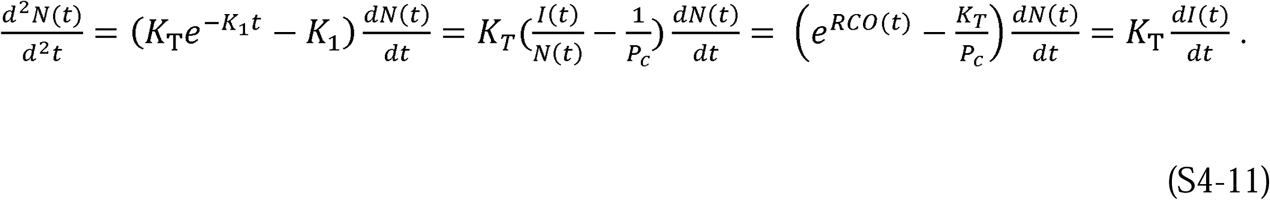

Equation S4-11, with its four equivalent expressions, demonstrates the power that an authentic model provides. The leftmost expression allows us to compare the acceleration—the potential to change the rate of new infections—at any stage of the epidemic for any two countries, even those with different population densities, using only the defining constants, *K*_*T*_ and *P*_*c*_, plus the daily case rate. Equation S4-11 is an immediate determinant of whether the control measures in place, represented by *P*_*c*_, are effective enough. If the value of the term 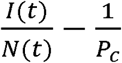 is positive, then the control measures are not strong enough. Conversely, when this term is negative, the epidemic is being brought under control.

The furthest right-hand equality in Equation S4-11, 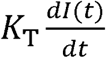, underscores that the goal of all containment measures should be to limit the creation of new infections as quickly as possible. When there are more daily new infections than daily recoveries, the epidemic is accelerating and expanding. Conversely, when there are fewer daily new infections than daily recoveries, the epidemic is slowing and will end, provided the containment measures are kept in place long enough to extinguish the outbreak.

The maximum value of *P*_*c*_ that will begin to bring down the new cases per day occurs when the acceleration is less than zero. If we set the left-hand side of Equation S4-11 to zero, use the third expression from the left and solve for *P*_*c*_, we arrive at the defining relationship for this critical parameter of epidemic management:

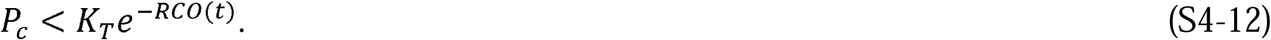

Since we can easily determine the value of *RCO*(*t*) every day during the epidemic and the value of *K*_*T*_ can be determined using the technique illustrated in Supplement 2, the maximum allowable value of *P*_*c*_ needed to reduce the number of daily cases can always be determined. This value of *P*_*c*_ is the maximum level of infectable social contact allowable if we want the number of new daily cases to continue decreasing.

Yet another important relationship can be derived from Equation S1-38. In that equation, the term 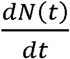 is the rate of new cases and, in Figures 3, 6B, D, F, H, 7, 9 and 11, this is the new cases per day. If we define a desired target for the number of new cases per day at a future time, *t* + *t*_*target*_, then we can define a new quantity, the desired fraction of the current new cases, *D*_*tf*_, as:

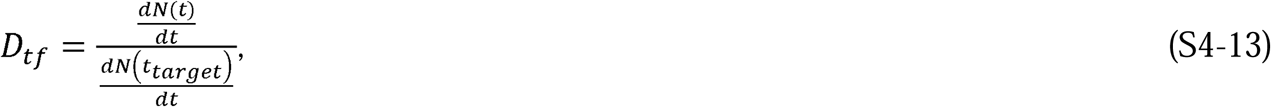

and using Equations S1-38 and S1-39, we arrive at the following expression:

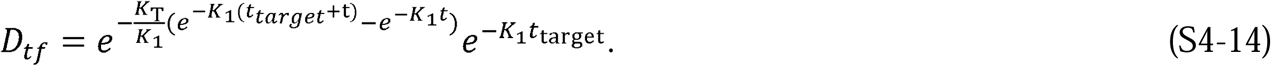

If *t* ≫ *t*_*target*_, then 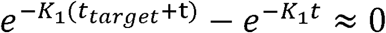 and we can derive the following equation from the remaining terms:

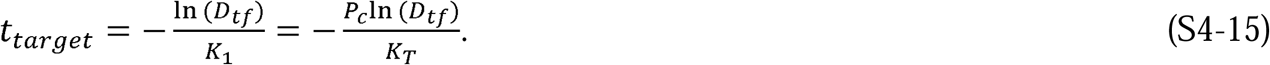

Equation S4-15 quantitates the number of days, *t*_*target*_, that a level of social containment, *P*_*c*_, must be imposed to achieve a fraction of daily cases, *D*_*tf*_, compared to the current level.

As a final comment, Equation S2-4 shows that, under the initial assumptions of immunity, no cases from outside, and a contiguous epidemic, *K*_1_(*t*) > 0 (See Supplement 7 for an explanation of contiguousness). Since *K*_1_(*t*) is inversely proportional to *A*_1_(*t*) and, as people reduce the area they traverse per unit time by increasing social distancing or taking other measures, *K*_1_(*t*) increases due to the lowered contact rate. Therefore, as formulated and as long as *K*_*T*_(*t*) is constant, *K*_1_(*t*) is inversely proportional to the strength of social distancing interventions implemented during an epidemic.

### Supplement 5. Controlling epidemics early

The quantitative mathematical relationships derived from the CSIR solution in Supplements 2 and 4 characterize the dynamics of an epidemic and illustrate that strong and early intervention is critical. Equation S4-1 quantifies that the ultimate number of individuals infected in an epidemic, *N*_∞_, will be exponentially dependent on the number of people with whom each person interacts.

The real-world country data provide vivid examples. Both South Korea and New Zealand enacted strong and early interventions compared to other countries (3, 4), as reflected by their *K*_*1*_ values (Table 1). These strong interventions led to earlier peaks in new cases and to far fewer total cases than in other countries (Figures 2 and 3): the peak number of new cases in both South Korea and New Zealand was 90–99% lower than in other countries, a compelling validation of the explicit statement in the CSIR solution that strong intervention leads to *exponentially* more favorable outcomes.

In the USA, interventions that began on March 16 started to have an effect around March 23, 2020 (Figure 3B); the number of active cases on March 23, 2020 (6) was 46,136 (Table 1). Using the values of *K*_*1*_ and *K*_*2*_ from Table 1, Equation S4-1 predicts that the ultimate number of cases would have been approximately 1.22 million. If the same intervention had been implemented and sustained starting on March 10, when there were 59 times fewer (782) cases (6), the model predicts that the ultimate number of cases would also have been 59 times lower, or 20,725. Thus, earlier action could have reduced the ultimate number of projected cases by more than 98%. Of course, the projected estimate of approximately 1.22 million total USA cases would only have occurred if the effectiveness of the interventions that were launched on March 16 had been sustained. Unfortunately, a marked reduction in effective interventions occurred in many parts of the USA in mid-April, well before the official reopening of the economy (13). This caused a second surge in new cases in late April and is why the observed data and the model prediction diverge in Figure 3B.

As shown in Supplement 4, the CSIR solution provides an estimate of the time to the peak of new cases, *t*_max_. Using Equation S3-10 and the values of *K*_*1*_ and *K*_*2*_ from Table 1, the predicted peak in new cases in the USA would have occurred near March 24 if the intervention had begun on March 10. Instead, a 6-day delay in effective intervention shifted the initial peak to April 11, 16 days later, as projected, and that peak was much higher (Figure 3B).

As shown, too, in Supplement 4, epidemic acceleration, the instantaneous potential to change the pace of the epidemic, can be determined at any point in the epidemic and depends on the social containment actions in effect at that time (Equation S4-11). What is perhaps less apparent, but predicted by the model, is that two countries with identical numbers of cases on a given day can, in fact, have different accelerations on the same day, and exhibit different dynamics immediately after that day.

South Korea and New Zealand (Figure 2A and F) had nearly identical case counts when each imposed strong containment measures (204 cases in South Korea on February 21, and 205 in New Zealand on March 25). Their models suggest that their interventions were about equally effective (*K*_*1*_ = 0.24 in South Korea and 0.17 in New Zealand; see Table 1). However, since South Korea has a much higher population density than New Zealand ((7), data in Table 2), it had a much higher number of interactions when the interventions were imposed and, therefore, a higher rate of acceleration, as evidenced by its higher RCO at the time of intervention. Indeed, the rate of change of new cases *was* higher in South Korea than in New Zealand, and the later number of cases in South Korea *was* higher than in New Zealand (Figure 2A and 2F).

Equation S4-8 clearly illustrates these lessons. As social distancing is strengthened (lower *P*_*c*_ and therefore, higher *K*_1_), the Effective Replication Number decreases, and the epidemic slows. Early and strong interventions, especially in countries with indigenously high levels of social interaction, are necessary to stop an epidemic in the initial stages. Re-openings, enacted too early, can reignite the epidemic, dramatically increasing the number of cases. The astonishing magnitude of the effects, driven by only a few days of delay, derives from the doubly exponential nature of the underlying relationships.

### Supplement 6. Ending an ongoing epidemic

We can use the CSIR solution to design measures to end an epidemic in an advanced stage. The management plan is built by first using Equation S4-13 to predict the number of days a given level of intervention, *K*_1_, is needed to reduce the new daily cases by a target fraction:

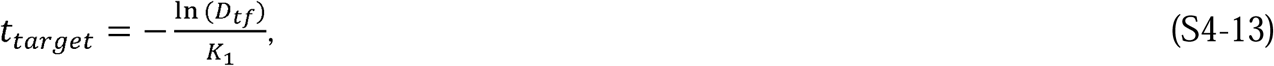

where *t*_*target*_ is the time to the desired reduction and *D*_*tf*_ is the target as a fraction of the current level of new cases per day.

For example, a country targeting a 90% reduction of new cases per day (e.g., from 50,000 to 5,000 cases per day, *D*_*tf*_ = 0.1), can attain its target in about 12 days by imposing a containment level of *K*_1_ = 0.2. The South Korea and New Zealand data demonstrate that Equation S4-15 is valid and that *K*_1_ = 0.2 is achievable for this duration. Both countries achieved a value of *K*_1_ close to 0.2 for the time necessary to produce a 90% reduction. It took 13 days in South Korea (March 3–16) and 15 days in New Zealand (April 2–15), New Zealand (6).

Since 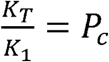, *K*_1_ = 0.2 characterizes a lockdown in which people in a country can each have only one plausibly infectious contact with a little over one specific person for the containment duration. This does not mean they cannot contact anyone other than the one person; but they must use care, masks and proper distancing, to ensure there is no plausibly infectious contact with anyone other than the one person.

Returning to the planning example, after achieving the initial 90% reduction, a reasonable next step might be to relax social containment to a level that allows the economy to remain viable, while preventing the epidemic from erupting again. We can again find the level of *K*_1_ necessary to achieve a chosen target, using Equation S4-13. If an additional 90% reduction in new cases per day is desired, and a period of 90 days is tolerable for that reduction, then a new level of approximately *K*_1_ = 0.025 is needed. This equates to a 90-day period during which each person can be in contact with seven specific people, in an infectable way. Note that this is three times *less* stringent than the original USA shutdown level in April 2020 as shown by the level of *K*_1_ calculated for the United States in that period (Table1). Thus, with a well-planned approach, a country can reduce its new daily cases by 99% in approximately 100 days, enabling the country to control, and essentially end the epidemic, while simultaneously maintaining economic viability.

If even *K*_1_ = 0.025 is too restrictive, we can choose a still lower *K*_1_, but it must be large enough to avoid a new outbreak. A bound for the new value of *K*_1_, low enough to prevent an outbreak and yet continue decreasing the new cases per day, can be found using Equation S4-12.

We can easily monitor the progress of interventions using the RCO, as the curve for South Korea illustrates (Figure 1A). Had this country maintained the implemented level of distancing measures, the data would have followed the initial slope. However, the actual data departed from the slope, heralding failures in (or relaxation of) social distancing, which were later documented to have occurred during the indicated time frame (3) (circled data, Figure 1A). Because it summarizes epidemic dynamics, we can use the RCO to continuously determine the effectiveness of implemented measures and whether they need adjustment.

#### S6.1 Outbreaks

Thus far in the development of the CSIR solution, we have assumed *K*_1_ to be constant over time. The solution is readily extended to allow *K*_1_ to vary with time in a piecewise manner. Here, we develop equations describing the analogous model in which *K*_1_ is constant in intervals of time, between which it changes. In addition, because this is a common happenstance, we develop an expression for *K*_2_ for use in Equation S1-39 when the model is fit to data that does not start at time *t* = 0.

If *K*_1_ and *K*_2_ are functions of time, Equation S1-38 can be rewritten as

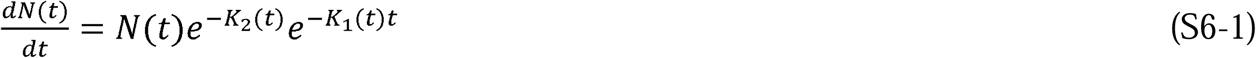

and Equation S1-39 as

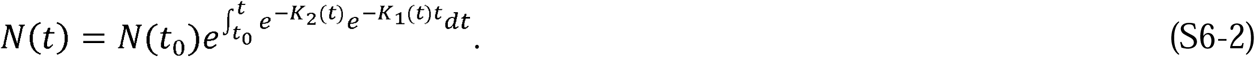

Distancing measures tend to be constant for many days at a time, so for this analysis, we assume *K*_1_ is piecewise linear. Therefore, we can calculate *K*_2_(*t*) for any time, *t*(*n*), when *K*_1_ changes:

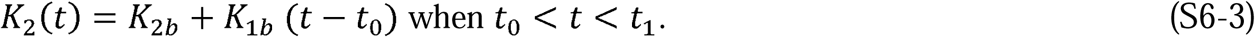

If *t*_*0*_ = 0, then *K*_1*b*_ and *K*_2*b*_ are the baseline levels of these parameters, and 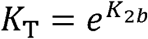. These three *K* values represent epidemic dynamics during the initial stages, before any containment measures are implemented.

As time passes, *K*_2_ becomes

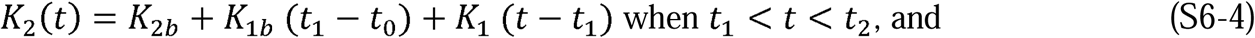

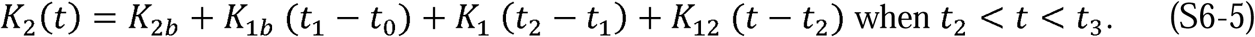

Noting the pattern as *t* increases, we can rewrite Equations S6-4 and S6-5 as

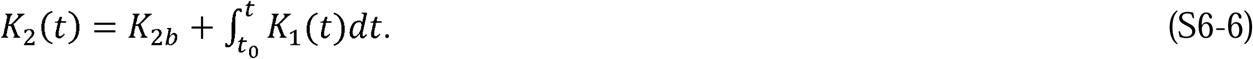

For any interval, *t*_*n*_ < *t* < *t*_*n*+1_, where *K*_1_ is a constant, *K*_2_(*t*) can be expressed as

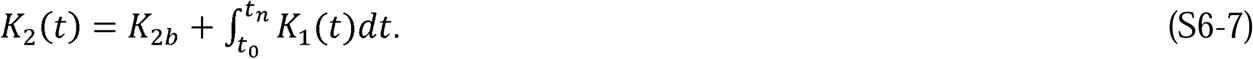

Equations S6-1 and S6-2 can also be rewritten as

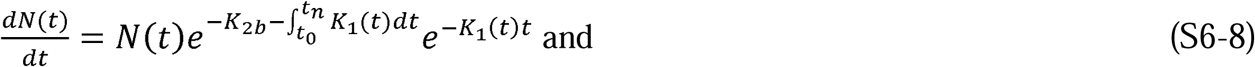

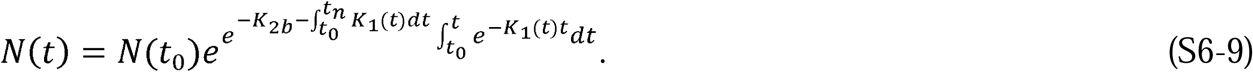

An expression for what happens in an epidemic *before* and *after* implementing social distancing measures is especially useful. A typical, or perhaps worst-case scenario might be a reopening of the economy, at which time social distancing measures are withdrawn and social contact returns to normal levels. In that instance, when distancing measures are lifted *K*_1_(*t*) becomes *K*_1*b*_ at time *t*_n_. Using Equation S1-38, we arrive at the following expressions:

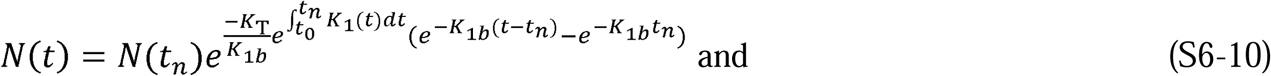

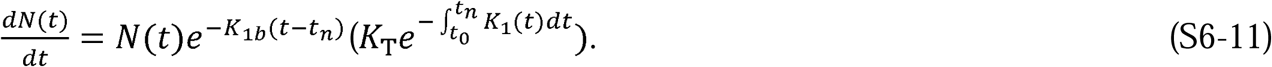

Equations S6-10 and S6-11 predict the progression of the epidemic before and after instituting interventions. They demonstrate that the dynamics of the epidemic depends on prior containment measures, as shown by the appearance of the expression 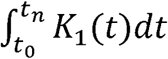 in both equations. This means that as long as the initial three assumptions of 1) immunity, 2) no new infections are introduced from outside the area, and 3) the epidemic remains contiguous (see Supplement 7) are still valid, then when interventions are relaxed, the epidemic can still grow nearly exponentially, but growth will be less rapid than the initial outbreak. This is a consequence of the fact that, under the three assumptions, *K*_1_ > 0. Since *K*_1_ is proportional to the inverse of the effective area traversed by an individual, it is proportional to the number of social interactions, and social interactions can never be less than zero. We label an outbreak when the three assumptions remain valid, but *K*_1_ decreases due to more social interaction, a Type 1 outbreak. A Type 1 outbreak appears to have occurred in mid-April in the USA because the slope of the RCO curve became less negative (Figure 1B).

If new infections are introduced into a portion of the population that has been thus far disconnected from the previously infected area, then the assumption of contiguity is violated. This is a common situation when infected people travel from an infected area to an area that was previously uninfected or had not yet seen significant numbers of infections. We label this a Type 2 outbreak.

Equation S1-39 must be modified to predict the number of cases in an epidemic affected by Type 2 outbreaks. Assuming that *t*_*0*_ = 0, *N*(*t*_0_) = 1, and introducing the notation *K*_1*x*_, where *x* denotes the number of the outbreak, Equation S1-39 can be written as:

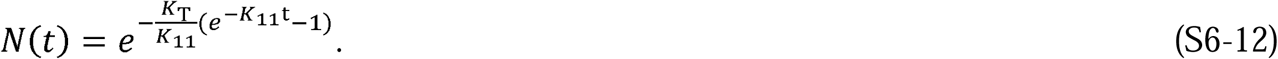

If a new outbreak occurs in a previously unaffected area of a country, then Equation S1-39 can be modified as follows:

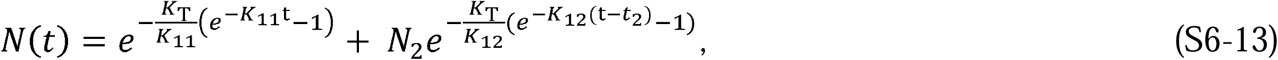

where *N*_2_ is the number of infectious people who initiated the new outbreak, *K*_12_ is the social interaction parameter in the new outbreak area, and *t*_*2*_ is the time the new outbreak occurs.

Equation S6-13 can be written in a general form as

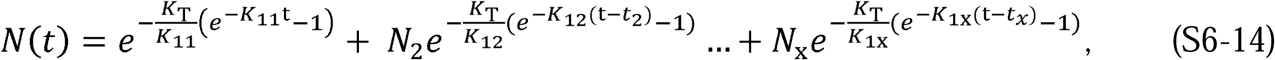

where *x* denotes the outbreak number and *t* > *t*_2_ > *t*_3_ > … > *t*_*x*_. For each outbreak *t*_*x*_, *K*_1*x*_, and *N*_x_ need to be determined independently.

While an epidemic is underway, we can detect a Type 2 outbreak by monitoring the slope of the RCO curve. A positive slope (*K*_1_ > 0) detected in an RCO curve indicates that a Type 2 outbreak has occurred. This is an indication that immediate action, within days, is required from policy makers to strengthen intervention measures and prevent the outbreak from overwhelming prior progress in controlling the epidemic.

By monitoring the RCO curve, we can also detect if the disease changes its transmissibility through mutation. In this situation, a proper fit of the parameters in Equation S1-44 is not possible and a modification of *K*_*T*_ is required to accommodate the change.

### Supplement 7: Understanding the assumption of contiguousness

To enhance the understanding of the CSIR solution and explain what the assumption of a contiguous epidemic means, we applied a perspective derived from the expression for *N*_∞_ in Equation S4-1 to Equations S1-1 to S1-5. Rather than look at the epidemic as involving the total population, *N*_*p*_, this perspective focuses on only that portion of the population that will eventually become infected in the epidemic. This is the subpopulation *N*_∞_. We also introduce a subpopulation of *N*_∞_ called *N*_*S*_(*t*), which we define as the sub-population at time *t* that is in contact with the epidemic and under threat of infection. This change in perspective views the epidemic mathematically from inside the bounds of the already infected population, which is ever expanding, rather than mathematically describing what is happening within a fixed total population.

The reinterpretation begins by first recognizing that, in Equations S1-1 to S1-3, the initial number of infections introduced to the population—which we designate *I*_*i*_—is merely the starting value of the epidemic and can be any value at all. The second step is to imagine that at every time, *t*, the epidemic starts again and the initial infections introduced into the population, *I*_*i*_(*t*), are equal to the then-current infections. The third step is to recognize that the values of the integrals in Equations S1-3 to S1-5 are equal to zero and *B*(*θ*) =1 whenever the epidemic starts. With these perspectives in mind, if Equations S1-1 to S1-3 and S1-5 are first all multiplied by *A*_*P*_, they can be rewritten as

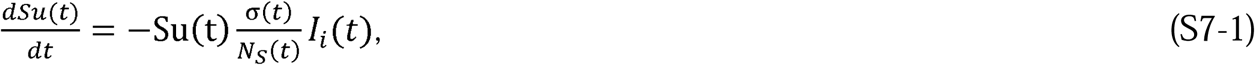

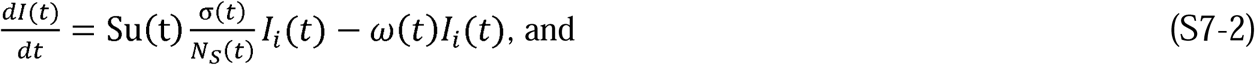

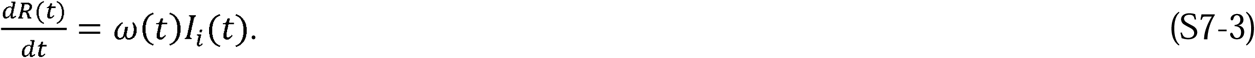

where *σ*(*t*) = *φ*(*t*) *N*_*S*_(*t*) and *ω*(*t*) = *ψ*(*t*),

*Su*(*t*) is the remaining portion of the population that will become infected, and since there are only susceptible people in *N*_*S*_(*t*) when the new infections are introduced,

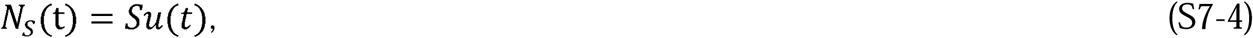

Of course, 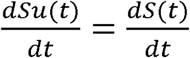.

If we also now define a quantity, *R*_*i*_(*t*), as the number of recovered persons introduced to the population at the same time as *I*_*i*_(*t*), we can write an equation for *N*_∞_ and define a new quantity, *N*(*t*), as the number of people either currently infected or recovered:

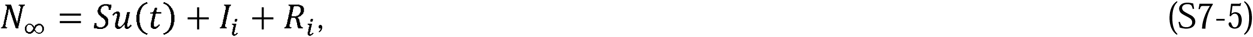

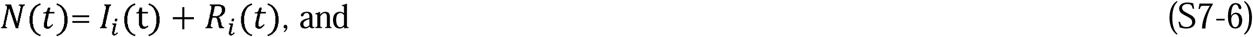

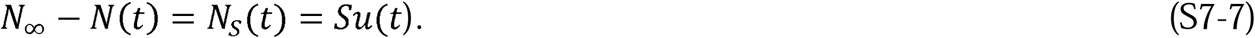

We have not assumed that any of the quantities are constant; therefore, we can now explicate the time-varying nature of these quantities. We begin by considering what happens to Equations S7-1 to S7-3 during the time interval Δ*t* from a time *t* to *t* +Δ *t*, and by rewriting these equations as difference equations:

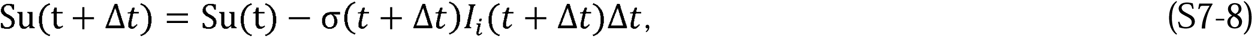

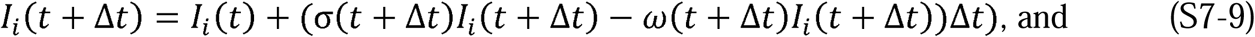

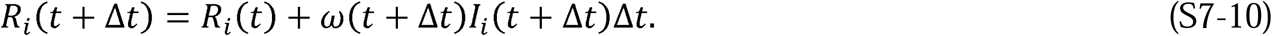

Taking the limit as Δ*t* → 0, we obtain the following differential equations:

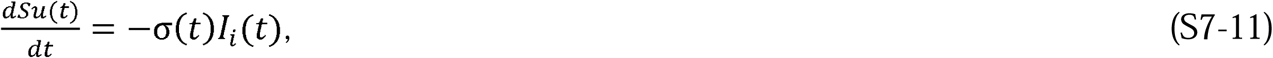

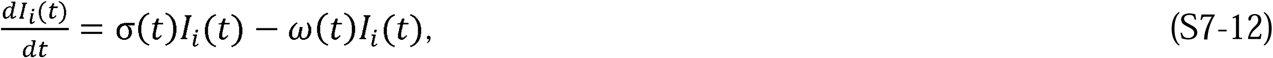

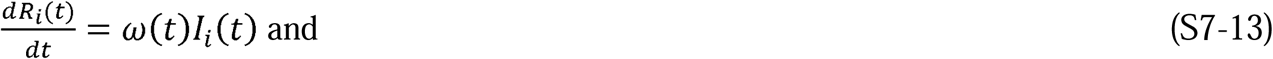

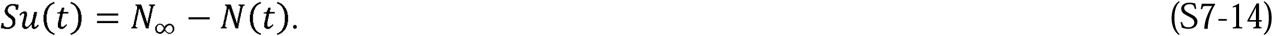

From the preceding, it can be easily seen that *I*_*i*_(*t*) = *I*(*t*) and *R*_*i*_(*t*) = *R*(*t*). Note that *Su*(∞) = 0 unlike *S*(∞).

The perspective in the immediately preceding part of the analysis is that the epidemic can be considered to start over again at each instant in time. In this perspective, the susceptible population *Su*(*t*) is not fixed by initial conditions, but rather is the population that will eventually become infected. Embedded in this concept is the assumption that during each Δ*t*, the susceptible population is always in contact with those people who have been previously infected or who will become infected. That is, the epidemic remains contiguous. This is not restrictive when considering the initial stage of the epidemic; however, as explained in Supplement 6, it plays a critical role in recognizing an outbreak that starts outside the population *N*_∞_.

We have recast the equations by shifting the population and area under consideration to *N*_∞_ and *A*_∞_, respectively, rather than *N*_*p*_ and *A*_*p*_. Further, we have defined the susceptible portion of the population during the epidemic as those who will eventually become infected under the conditions in place at each instance in time. This shift in perspective retains the mathematical equivalence to the equations derived by Kermack and McKendrick (1) because the portion of *N*_*p*_ that is not a part of *N*_∞_ never becomes infected, and therefore never affects the values of *I*(*t*) or *R*(*t*) in the solutions to Equations S1-1 to S1-5 or S1-8 to S1-11.

